# Dopaminergic sub-network connectivity alterations are associated with postoperative cognitive dysfunction: Results from the BioCog cohort study

**DOI:** 10.1101/2025.01.02.25319918

**Authors:** Florian Lammers-Lietz, Friedrich Borchers, Insa Feinkohl, Cicek Kanar, Henning Krampe, Gregor Lichtner, Jayanth Sreekanth, Janine Wiebach, Martin Weygandt, Claudia Spies, Georg Winterer, Friedemann Paul, the BioCog consortium

## Abstract

Postoperative cognitive dysfunction (POCD) is a detrimental complication after surgery with lasting impact on the patients’ daily life. It is most common after postoperative delirium. Dopaminergic dysfunction has been suggested to play a role in delirium, but little knowledge exists regarding its relevance for POCD. We hypothesized that POCD is associated with altered resting-state functional connectivity of the ventral tegmental area (VTA) and the substantia nigra pars compacta (SNc) in functional magnetic resonance imaging (fMRI) before surgery and at postoperative follow-up after three months.

Patients ≥65 years old underwent resting-state fMRI and neuropsychological assessment before major elective surgery and at follow-up three months later. POCD was determined as the reliable change index. Connectivity between VTA or SNc and 132 regions were calculated. Principal component analysis (PCA) was used for sub-network construction, and components explaining >5% of variance were retained for analysis. To study postoperative changes in patients with POCD, we applied the same transformation to postoperative connectivity, and multi-factor analysis. Regression analyses were used to describe connectivity alterations while adjusting for age, sex, MMSE, surgery and anaesthesia.

Of 214 patients, 26 (12%) developed POCD. Among 132 principal components, four components for VTA- and SNc-FC were selected for further analysis. For both VTA and SNc connectivity, one component was significantly associated with POCD. Postoperative alterations of dopaminergic networks were observed in an exploratory voxel-wise analysis in a left temporal cluster.

Higher dopaminergic connectivity to regions associated with spatial perceptive processes and lower connectivity to cognitive control-related areas may predispose to POCD.

**SIGNIFICANCE STATEMENT:** Delirium affects around 20% of older adults after surgery, with 30% developing lasting memory and attention issues (postoperative cognitive dysfunction, or POCD). While antidopaminergic drugs are used to treat delirium, they do not prevent long-term problems. To explore dopamine’s role in cognitive issues after delirium, we used MRI scans before and after surgery. We found that in patients who later developed POCD, dopamine-related brain regions communicated differently before surgery compared to those who did not develop POCD. These changes appear to be chronic, meaning delirium treatments may have little effect on long-term cognitive problems.

## 1 Introduction

Lasting postoperative cognitive dysfunction (POCD) affects 10-50% of older patients undergoing surgery (Androsova *et al*. 2015; Borchers *et al*. 2021). Especially patients who experienced postoperative delirium (POD) have a significantly increased risk (Rudolph *et al*. 2008; Teipel *et al*. 2018). POCD is associated with increased mortality, length of hospital stay (Suraarunsumrit *et al*. 2024), care costs (Pietzsch *et al*. 2021) and labor market leave (Steinmetz *et al*. 2009).

Dopaminergic dysfunction is thought to be a hallmark of POD (Maldonado 2013; Androsova *et al*. 2015), as suggested by genetic studies (van Munster *et al*. 2010b, 2010a) and clinical observations from prodopaminergic (Kuzuhara 2001; Schrag 2004; Mash 2016; Gonin *et al*. 2018) and antidopaminergic substances (Aldecoa *et al*. 2024). However, there is no evidence that antidopaminergic treatment of POD has a beneficial effect on long-term cognitive outcomes: Whereas it is well known that shorter delirium duration is associated with more favorable long-term outcomes (Pisani *et al*. 2009) including cognition (Girard *et al*. 2010), the perioperative application of antipsychotic medication was associated with postoperative cognitive decline (Duprey *et al*. 2022). In the past, two studies have used functional magnetic resonance imaging (fMRI) to describe ventral tegmental area (VTA) connectivity changes in POD using fMRI, but comparable works for POCD are lacking (Choi *et al*. 2012; Oh *et al*. 2019)

Several lines of evidence, including fMRI approaches, found alterations of the dopaminergic system in various neurodegenerative diseases (Holmes *et al*. 2001; Craig *et al*. 2004; Sweet *et al*. 2005; Serra *et al*. 2018, 2021; Li *et al*. 2020; Yoo *et al*. 2022; Zang *et al*. 2022), suggesting that dopaminergic dysfunction could account for the lasting cognitive symptoms in POCD. An investigation on the dopaminergic system in POCD could also help to clarify the causal relationships between POD and POCD: Since POD is more common in patients with preoperative cognitive dysfunction (Dasgupta and Dumbrell 2006; Androsova *et al*. 2015), POCD may also reflect manifestation of a preoperative dopaminergic dysfunction.

On the other hand, dopaminergic activity is a driver of emergence from general anaesthesia (Solt *et al*. 2014; Zhou *et al*. 2015). While dopaminergic neurons show tonic background activity, (Paladini and Roeper 2014), it has been found that under cortical slow wave activity during general anaesthesia, and in response to painful stimuli, dopaminergic activity increases (Walczak and Błasiak 2017; Izowit *et al*. 2024). While this may cause hyperdopaminergic states in POD, it could also pose extraordinary stress to dopaminergic neurons during anaesthesia and surgery. A variety of radicals is generated during dopamine synthesis as well as degradation, and especially a high turn-over of dopamine, e.g., during anaesthesia and POD, or as part of a sympathetic response to surgery, may elicit oxidative stress to dopaminergic neurons and trigger cell death (Beal 1995; Schmidt and Reith 2005; Labandeira-García *et al*. 2014). Oxidative stress in dopaminergic neurons may particularly occur under circumstances of impaired cellular energy metabolism as a result of excitotoxicity, and in addition, the tonic baseline activity of dopaminergic neurons may render the particularly vulnerable to hypoperfusion and ischaemia (Beal 1995; Schmidt and Reith 2005; Paladini and Roeper 2014). These conditions occur regularly during anaesthesia and surgery, e.g., due to blood loss, ventilatory impairment, and direct effects of anaesthetics (Berthoud and Reilly 1992), that are known risk factors for POCD (Aceto *et al*. 2020; Kitthanyateerakul, Tankumpuan and Davidson 2024; Djurasovic *et al*. 2025).

In conclusion, existing literature suggest a complex interplay between surgery, dopaminergic function and cell loss and POCD. Hence, there is a need to understand the longitudinal relationship between the dopaminergic functiona and POCD.

The central nervous dopaminergic system comprises the substantia nigra pars compacta (SNc, A9), containing 75-90% of all dopaminergic neurons in the central nervous system. The SNc has reciprocal connections with the striatum, with essential function for motor control and cognition (Mai and Paxinos 2011). About 50% of the neurons in the VTA (A10) are dopaminergic. The VTA projects to nucleus accumbens, hypothalamus, amygdala, hippocampus, the cortex and has reciprocal connections with noradrenergic, serotonergic, cholinergic and GABAergic brainstem regions, as well as connections to the nucleus accumbens and globus pallidus, limbic and neocortical areas (Mai and Paxinos 2011).

In fMRI, correlations of fluctuations in the blood oxygen level-dependent (BOLD) signal can be used to derive measures of co-activation between anatomically distinct brain regions, i.e., functional connectivity. Functional connectivity patterns under rest are referred to as resting state (rs) networks, and rs-fMRI of the dopaminergic system could provide helpful information on dopaminergic dysconnectivity in POCD, but to our knowledge, no such study has been conducted.

Here, we present analyses of fMRI data from the BioCog study, a cohort study targeting the prediction of POD and POCD in cognitively healthy female and male patients ≥65 years undergoing elective surgery. We hypothesized that VTA and SNc functional connectivity alterations predisposes to POCD, and furthermore, that alterations would parallel the development of POCD.

We calculated functional connectivity of the VTA and SNc with 132 other cortical and subcortical brain regions before surgery and at follow-up three months after surgery. To account for the diffuse projection patterns of the dopaminergic system, we then used principal component analysis (PCA) to construct dopaminergic functional sub-networks, and analyzed the association of POCD with selected VTA and SNc subnetworks.

## 2 Methods

### 2.1 Study design

The BioCog (Biomarker Development for Postoperative Cognitive Impairment in the Elderly, www.biocog.eu; clinicaltrials.gov: NCT02265263) study is an EU-funded multicenter prospective observational cohort study with the aim of identifying risk factors for POD and POCD. Patients were enrolled at the Charité – Universitätsmedizin Berlin, Germany, and the University Medical Center Utrecht, Netherlands. The study was conducted in line with the Helsinki Declaration. All procedures were approved by the local ethics committees in Berlin, Germany (EA2/092/14) and Utrecht, Netherlands (14-469).

### 2.2 Participants

Here, we analyse a subsample of 214 surgical patients from the Berlin cohort of the BioCog study. Consenting patients aged ≥65 years presenting for elective surgery with an expected duration >60min were included. Patients were excluded from study in case of positive screening for pre-existing major neurocognitive disorder defined as a Mini-Mental Status Examination (MMSE) score ≤23 points, any condition interfering with neurocognitive assessment (severe sensory impairment, neuropsychiatric illness), expected unavailability for follow-up assessment, accommodation in an institution due to official or judicial order or inability to give informed consent. All participants gave written informed consent prior to inclusion.

### 2.3 Outcome: POCD

Neuropsychological testing (CANTAB test battery, RRID:SCR_003001, Trail-Making-Test pt. B, and the Grooved Pegboard Test) took place at baseline before surgery, and three months after surgery. To adjust for natural variability in cognitive performance and learning effects in repeated cognitive testing, POCD was defined according to the Reliable Change Index (RCI) with the ISPOCD criteria proposed by Rasmussen (Rasmussen *et al*. 2001) implemented in the POCD package for R (https://github.com/Wiebachj/POCDr) (Spies *et al*. 2021). The RCI is the patient’s change in performance at a cognitive test parameter after surgery compared to baseline performance in relation to the mean change in a non-surgical control group. POCD was diagnosed in patients with an RCI score below -1.96 in at least two single cognitive test parameters (i.e., showing a more severe performance decline than approximately 97.5% of the reference population) and/or a compound RCI averaged over all single RCIs in the follow-up assessment three months after surgery. A more detailed description is provided in previous publications (Lammers *et al*. 2018; Feinkohl *et al*. 2020; Spies *et al*. 2021) and in the supplement.

### 2.4 Neuroimaging

#### 2.4.1 MRI acquisition

MRI data were collected before surgery at the Berlin Center for Advanced Neuroimaging using a 3T Magnetom Trio MR scanner (Siemens, Erlangen, Germany). After three months, patients were invited to a follow-up session at the same scanner.

Resting-state fMRI was performed using a 12-channel head coil and 2D echo-planar blood oxygenation level dependent (BOLD) sequence (7.8min duration, 238 slices, field of view: 192·192 mm in 32 transverse slices, 3·3·3mm³ isotropic voxels, repetition time=2.0s, excitation time=30ms, descending slice order, 78° flip angle). Resting-state fMRI was conducted at second position of the whole MRI protocol and was completed approximately 15min after start of the whole session to minimise the risk of the patients falling asleep. Immediately before start of the sequence, the patients were contacted verbally to ascertain that they were awake before the start of the scan. The scanning room was darkened and the patients were instructed to close their eyes and to rest quietly without mental focusing in the scanner bore.

T1-weighted 3D anatomical brain scans were acquired with a 32-channel head coil using a magnetization prepared rapid gradient echo (field of view: 256·256 mm² in 192 sagittal slices, 1mm³ isotropic voxels, repetion time=2.5s, exitation time=4.77ms, 7° flip angle).

#### 2.4.2 Seed ROI definition

Seed ROIs of the substantia nigra, pars compacta (SNc, A9, 506mm³ volume) and ventral tegmental area (VTA, A10, 805mm³ volume) were defined by masks from the Brainstem Navigator 0.9 (Brainstem Imaging Lab, Athinoula A. Martinos Center for Biomedical Imaging) (Bianciardi *et al*. 2015, 2018; García-Gomar *et al*. 2019, 2022; Singh *et al*. 2019; Singh, García-Gomar and Bianciardi 2021; Brainstem Navigator 2021).

#### 2.4.3 Functional MRI preprocessing and connectivity analyses

Results included in this manuscript come from analyses performed using CONN (RRID:SCR_009550) 22.a and SPM 12.7771 (RRID:SCR_007037) for MATLAB R2022b (RRID:SCR_001622) (Penny *et al*. 2006; Whitfield-Gabrieli and Nieto-Castanon 2012; Nieto-Castanon and Whitfield-Gabrieli 2022). The default prepocessing and denoising pipeline implemented in CONN have been used.

ROI-to-ROI connectivity matrices and seed-based connectivity maps were estimated characterizing the patterns of functional connectivity between the two seed ROIs (VTA, SNc) and 132 target ROIs from the Harvard-Oxford atlas. Functional connectivity strength was represented by Fisher-transformed bivariate correlation coefficients (Nieto-Castanon and Whitfield-Gabrieli 2022), defined separately for each pair of seed and target areas, modeling the association between their BOLD signal time series. Additional details on the neuroimaging methods are given in the supplementary material.

### 2.5 Dimensionality reduction in resting-state functional connectivity data and construction of sub-networks

While voxelwise analysis is the most common approach in functional connectivity studies, we decided to use principal component analysis (PCA) and multi-factorial analysis (MFA) to reduce data dimensions in preoperative functional connectivity data for VTA and SNc and construct dopaminergic sub-networks from dopaminergic resting-state functional connectivity.

Dimensionality reduction by PCA seemed reasonable from a biological perspective, as outlined in the introduction. In contrast to GABA and glutamate, cerebral concentrations of dopamine are low. Furthermore, dopaminergic neurotransmission is terminated by uptake via dopamine and norepinephrine transporters (DAT and NET), which are not located in the synaptic cleft. Hence, dopamine molecules diffuse from the site of release and act on receptors in the distance before removal (volume transmission) (Brady *et al*. 2012). Dopaminergic neurons from the VTA innervate vast areas of the human and primate cortex (Berger, Gaspar and Verney 1991; Zubair *et al*. 2021). Hence, a process affecting dopaminergic neurotransmission might not exert strong, regionally limited, but rather subtle and widely distributed effects on BOLD signal fluctuations in sub-networks. Delirium is an encephalopathy as a consequence of systemic disturbances rather than an inherent brain disorder (Androsova *et al*. 2015; Maldonado 2018; Aldecoa *et al*. 2024) and delirium-related cerebral dysfunction, including POCD, are expected to affect the whole brain (Cavallari *et al*. 2016, 2017; van Montfort *et al*. 2018a, 2018b; Fislage *et al*. 2024). Hence, a transformation of the data allowing to select components related to global BOLD signal fluctuations over components to regionally limited signal changes may be a better aproximation to the exptected underlying neurobiology of POCD and dopaminergic network function.

In addition, previous research provided too little information for a hypothesis-driven analysis in a restricted set of target ROIs. However, the number of available functional connections was relatively high compared to the number of patients in the sample, and conducting analyses as independent tests was not possible without either significant loss of statistical power or an inacceptable increase in false-positive results. Last but not least, we expected high collinearity in the functional connectivity data, and PCA was used to transform them into sets of orthogonal variables for regression analysis.

However, to allow comparisons with studies using voxelwise analyses, we also provide results from voxelwise analyses for comparison.

Figure 1 provides an overview on the applied work-flows, covering derivation of functional connectivity measures, dimensionality reduction and statistical models.

**Figure 1:**
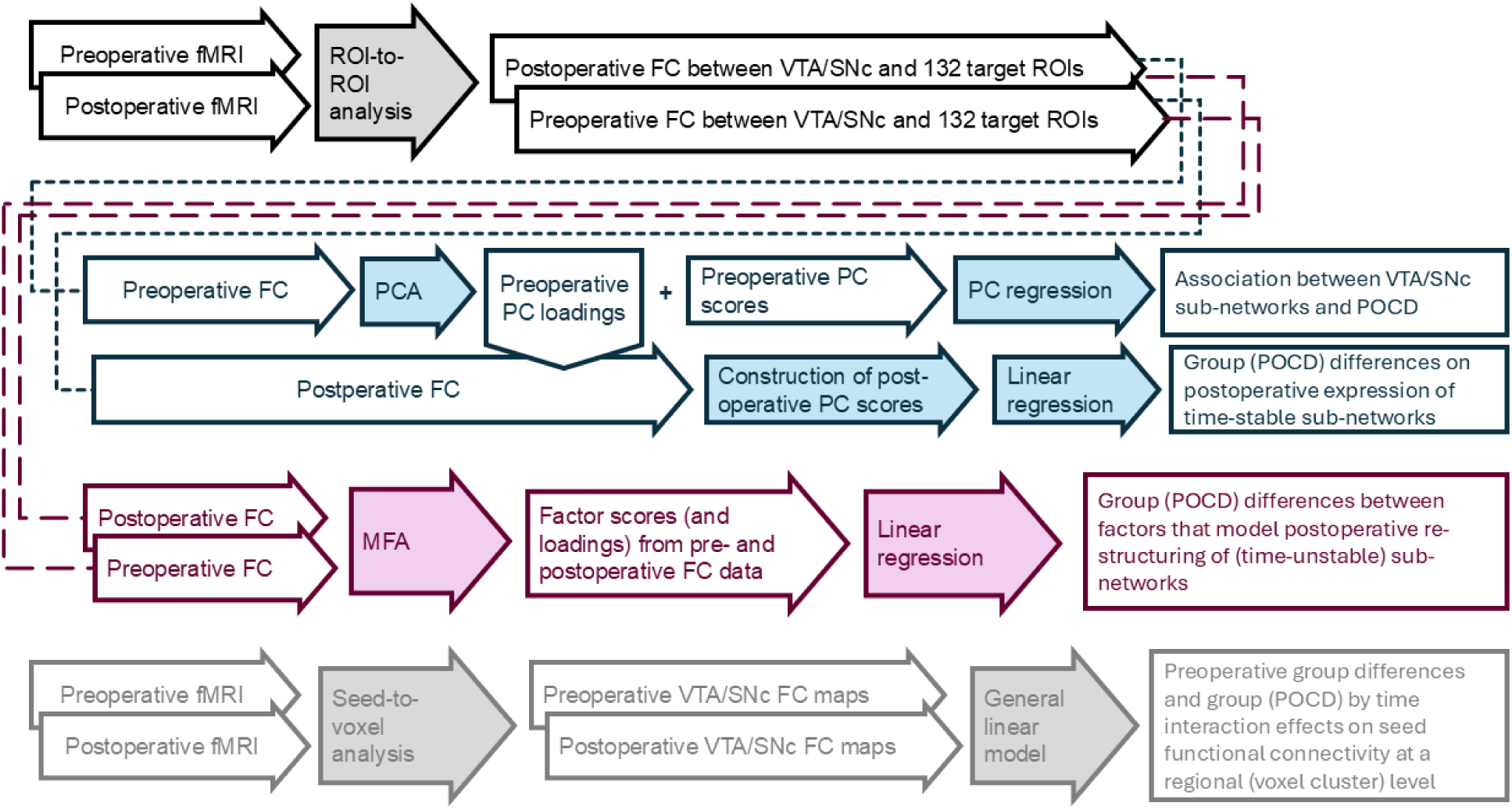
**Study work-flow.** Top: ROI-to-ROI functional connectivity matrices are derived from pre- and postoperative fMRI data that describe the functional connectivity between the seed ROI in the VTA or SNc, respectively, with 132 target ROIs. Depicted in blue: Principal component analysis (PCA) is conducted in preoperative FC data to derive VTA/SNc principal component reflecting preoperative inter-individual differences in sub-network expression. Principal components scores are used for principal component regression to analyse associations between VTA/SNc sub-network expression and development of POCD. PC loadings from preoperative data are also used to calculate postoperative component scores. In a linear regression model, group differences in postoperative, baseline-adjusted sub-network expression (i.e., postoperative component scores) between patients with POCD and control patients were assessed. Since in this approach, postoperative components are calculated from PC loadings of preoperative functional connectivity, a general assumption is that sub-networks are time-stable and not affected by time or surgery in general. Depicted in red: To consider effects of time or surgery on sub-networks construction, we conducted multi-factorial analysis (MFA) of pre- and postoperative ROI-to-ROI functional connectivity data. While MFA is generally similar to PCA, it was designed for analysis of longitudinal data. Factors from MFA reflect inter-individual differences in sub-network expression, but inherently describes temporal changes in (time-unstable) sub-networks and can be analysed in linear regression analysis for group differences between patients with POCD and control patients. Bottom: For comparison with sub-network focussed analyses, seed-to-voxel connectivity maps are calculated for VTA and SNc from pre- and postoperative fMRI data. Voxelwise analyses of VTA- and SNc FC in a general linear model describe preoperative regionally limited differences between patients with POCD and control patients, as well as the group by time interaction.

#### 2.5.1 Preoperative data

PCA was conducted separately in two data matrices of preoperative VTA and SNc functional connectivity separately. Matrix rows reflected patients and each matrix column reflected the functional connectivity of the seed region to one of 132 target ROI of the Harvard-Oxford atlas. While brain regions that show either extreme absolute functional connectivity values with the VTA or SNc seed regions are usually considered the VTA- or SNC-networks, PCA decomposes the functional connectivity between the seed regions and the target ROIs in mutually uncorrelated components. These components reflect subtle inter-individual differences in VTA- and SNc network patterns and composition and are expressed at distinct levels in each individual. They can be, in reverse, be used to re-construct the VTA- or SNc networks. It may hence be legitimate to refer to principal compontens as “sub-networks”.

Principal components explaining at least 5% of variance in the preoperative functional connectivity were considered for further analysis. This criterion was chosen post-hoc after inspection of PCA results, since we found that commonly used rules-of-thumb to determine the number of components to retain were not expedient (Keho 2012): Retaining all principal components with eigenvalues >1 or explaining well above 50% of the cumulative variance would still have resulted in an inacceptably high number of independent test, and the Cattell scree test was not conclusive. In addition, the amount of variance explained by each component was generally low, and we considered that retaining principal component explaining less than 5% of data variance would no longer fit the assumption on dopaminergic transmission outlined above.

#### 2.5.2 Longitudinal analyses

For longitudinal analyses of VTA and SNc functional connectivity in POCD, postoperative sub-networks were constructed by multiplying postoperative functional connectivity data with factor loadings from PCA of preoperative functional connectivity data (Brasanac *et al*. 2022; Lammers-Lietz *et al*. 2023). Since these scores were not derived from PCA directly, we refer to them as postoperative components rather than principal components. Step by step, PCA of preoperative functional connectivity data yields: X_pre_V_pre_=U_pre_ (X_pre_: matrix of original preoperative functional connectivity data; V_pre_: matrix with factor loadings derived from PCA; U_pre_: matrix with preoperative factor scores). Postoperative components U’_post_ were calculated as X_post_V_pre_=U’_post_ (X_post_: matrix with postoperative functional connectivity data).

We hypothesized that not only functional connectivity strength between seed and target ROI might be affected by surgery and change over time, but also sub-network composition might change over time, and the assumption that preoperative factor loadings apply to postoperative networks might not hold. In an additional approach, exploratory approach, we considered multi-factor analysis (MFA), which extends the concept of PCA to grouped or multi-table data, i.e., neuroimaging data collected at two distinct fMRI sessions (Abdi, Williams and Valentin 2013). MFA calculates a matrix of variable loadings of both pre- and postoperative data without a-priori assumptions on relations between variables (i.e., without assumption of identical networks over time) and then provides factor scores spanning both pre- and postoperative functional connectivity data. MFA was conducted separately for VTA and SNc connectivity in two separated data matrices with each row representing a patient with available pre- and postoperative fMRI data. Each of the 164 columns in each matrix represents a functional connection between the seed and one of 132 target ROIs from one of two fMRI sessions. Columns were weighted by fMRI session.

#### 2.5.3 Interpretation of sub-networks

To provide an interpretation of the sub-networks, we assigned functional labels to each preoperative principal component included in the regression analyses, using the NeuroQuery meta-analysis tool for human brain mapping (Dockès *et al*. 2020). NeuroQuery is intended to predict brain activation patterns from a list of key terms. To generate a brain map for a single query, the list of key terms is expanded by addition of weighted, related terms using semantic smoothing. NeuroQuery then uses the estimated semantic associations to map the query onto a set of keywords that can be reliably associated with brain regions. We took advantage of the literature database integrated in NeuroQuery and its semantic smoothing function to identify functions and conditions related to certain brain regions related to certain principal components.

Target ROIs with hight component loadings were entered into NeuroQuery, keywords identified by semantic smoothing for expansion of the meta-analysis were noted, and the associated publications referenced by NeuroQuery were reviewed. Lists of exact key terms entered in NeuroQuery as well as relevant publications referenced by NeuroQuery are described in the supplementary material.

### 2.6 Statistical analyses

#### 2.6.1 Considerations regarding statistical power

A power calculation for the primary aim of the BioCog study (i.e., development of a multivariable risk predictor for postoperative delirium) is given elsewhere (Lammers-Lietz *et al*. 2024). For this analysis, no separate power calculation was conducted, but all available datasets considered.

#### 2.6.2 Regression analyses of functional sub-networks

All statistical analyses were conducted in R (RRID:SCR_001905). 95% confidence intervals (CI) for regression coefficients (B) and odds ratios (OR) were calculated based on 100,000 bootstrap iterations.

##### 2.6.2.1 Preoperative data

Two separate logistic regression models were used to analyse associations of either preoperative VTA or SNc functional connectivity with POCD as the dependent variables. No occurrence of POCD was the reference category (assigned values: 1 for POCD, 0 for no POCD occurrence). Separate models for VTA and SNc functional connectivity were set up, yielding a total number of four regression models. For each seed ROI, all principal components explaining more than 5% of variance were included as the independent variables of interest in one single logistic regression model. All analyses were adjusted for age, sex, MMSE score at inclusion, ASA physical status, type of surgery and anaesthesia as well as duration of anaesthesia. The significance threshold was set to p<0.05 and Benjamini-Hochberg-adjustments were made for testing the total amount of principal components over both regression models.

##### 2.6.2.2 Longitudinal data

Postoperative components were tested for difference between patients with and without POCD three months after surgery in linear regression models. Postoperative components were treated as the dependent variables. POCD incidence was the independent variable of interest. All analyses were adjusted for preoperative principal component score, and interaction between preoperative principal component score and POCD, age, sex, MMSE score at inclusion, ASA physical status, type of surgery and anaesthesia as well as duration of anaesthesia.

Multi-factor analyses results were analysed as dependent variables in linear regression models with POCD as the independent variable of interest adjusted for age, sex, MMSE score at inclusion, ASA physical status, type of surgery and anesthesia as well as duration of anesthesia. Factors were not adjusted for preoperative scores since factors are derived from both pre- and postoperative functional connectivity and the regression models reflect group differences in perioperative dopaminergic network differences.

The level of significance was set to p<0.05 and adjusted for the total number of components considered in the longitudinal connectivity analysis using the Benjamini-Hochberg method.

#### 2.6.3 Voxel-level analyses

To facilitate comparison of our data with other studies using voxelwise analyses, and to address potential regionally restricted POCD-related alterations in dopaminergic connectivity, group-level analyses were performed using a General Linear Model (GLM) (Nieto-Castanon 2020). For each individual voxel a separate GLM was estimated, with first-level connectivity measures at this voxel as dependent variables (one independent sample per subject and one measurement per session), and POCD as the independent variable of interest adjusted for age, sex, MMSE score at inclusion, ASA physical status, type of surgery and anesthesia as well as duration of anesthesia. Voxel-level hypotheses were evaluated using multivariate parametric statistics with random-effects across subjects and sample covariance estimation across multiple measurements. Inferences were performed at the level of individual clusters (groups of contiguous voxels). Cluster-level inferences were based on parametric statistics from Gaussian Random Field theory (Worsley *et al*. 1996; Nieto-Castanon 2020). Results were thresholded using a combination of a cluster-forming p<0.001 voxel-level threshold, and a familywise corrected p-FDR<0.05 cluster-size threshold (Chumbley *et al*. 2010).

## 3 Results

### 3.1 Description of the cohort

5294 patients were screened and 747 patients were included in Berlin, of whom 61 dropped out before preoperative assessments. 320 patients enrolled at the Charité-Universitätsmedizin Berlin provided fMRI data. One subject was excluded prior to preprocessing due to a significant brain lesion as described earlier (Lammers *et al*. 2018). Another 13 patients were excluded during the quality assessment, leaving 306 patients in total for further analysis. 196 out of 306 patients underwent fMRI at follow-up after three months. 214/306 patients with preoperative data and 193/196 patients with pre- and postoperative fMRI data also provided POCD assessments. In total, 26 out of 214 patients (12%), and 24 out of 193 patients with MRI at follow-up (12%) had POCD. In total, 24 patients with fMRI at baseline and 20 patients with fMRI at follow-up had postoperative delirium.

Table 1 characterizes the cohorot of 214 patients with preoperative fMRI data and POCD assessment. The flow chart is presented in figure 2. Supplementary figure S1 compares the distribution of RCI between the sub-cohort presented here and control participants.

**Figure 2:**
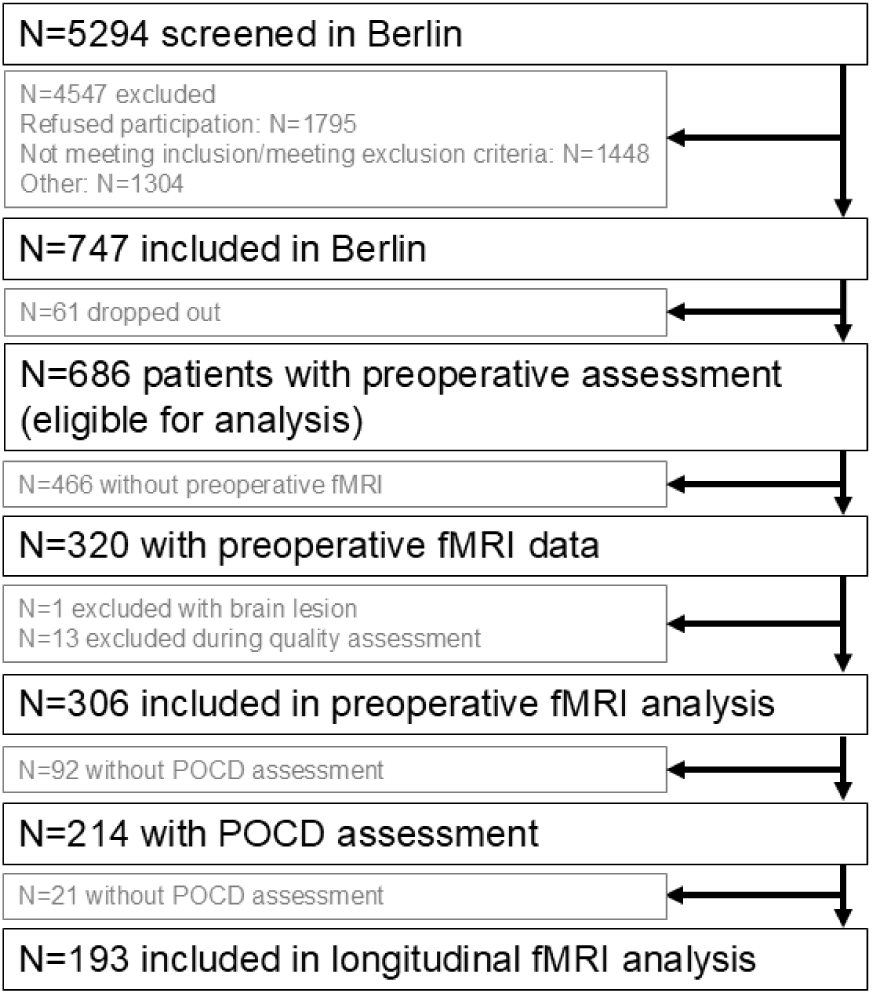
Patient flow chart. Among the 1448 screened patients who did not meet in- or exclusion criteria, 21 (0.4%) were excluded due to pre-existing dementia, 20 (0.4%) due to another neuropsychiatric disease possibly interfering with the neurocognitve testing and 30 (0.6%) were excluded due to an MMSE<24 points. 37 (0.7%) were excluded due to the inability to give informed consent or legal guardianship.

**Table 1:** Description of 214 patients with preoperative fMRI data.

| **TABLE 1: Description of 214 patients with preoperative fMRI data.** |  |  |  |
| --- | --- | --- | --- |
|  | **All** | **No POCD** | **POCD** |
|  | **N (%)** |  |  |
| POCD | 26 (12%) |  |  |
| POD | 24 (11%) | 21 (11%) | 3 (12%) |
| female | 86 (40%) | 75 (40%) | 11 (42%) |
| Intracranial surgery | 2 (1%) | 0 (0%) | 2 (8%) |
| Intrathoracal/-abdominal/-pelvic surgery | 73 (34%) | 66 (35%) | 7 (27%) |
| ISCED 1-2 (N=193) | 34 (18%) | 29 (17%, N=169) | 5 (21%, N=24) |
| ISCED 5-6 (N=193) | 82 (42%) | 73 (43%, N=169) | 9 (38%, N=24) |
| Regional | 10 (5%) | 9 (5%) | 1 (4%) |
| Combined general/regional | 35 (16%) | 33 (18%) | 2 (8%) |
|  | **Median, IQR**<br>**(Min-max.)** |  |  |
| Age (y) | 72, 68-75<br>(65-87) | 72, 68-75<br>(65-86) | 75, 70-79<br>(67-87) |
| MMSE (p) | 29, 28-30<br>(24-30) | 29, 28-30<br>(24-30) | 29, 28-30<br>(27-30) |
| ASA-PS | II, II-II<br>(I-III) | II, II-II<br>(I-III) | II, II-III<br>(II-III) |
| Duration of anaesthesia (min) | 143, 100-220<br>(36-506) | 143, 103-224<br>(36-506) | 142, 90-200<br>(45-359) |
| CCI (p) | 1, 0-2<br>(0-9) | 1, 0-2<br>(0-8) | 1, 0-2<br>(0-9) |
| *A deviating N is stated in case of missing data.*<br>*ASA-PS: American Society of Anesthesiologists Physical Status, CCI: Charlson comorbidity index, ISCED:*<br>*International Standard Classification of Education, MMSE: Mini-Mental Status Examination, POD: postoperative*<br>*delirium, POCD: postoperative cognition dysfunction;* |  |  |  |

### 3.2 Sub-networks

Supplementary figure S2 displays VTA and SNc resting-state functional networks as identified for the whole cohort from a voxelwise analysis. PCA of the ROI-to-ROI functional connectivity analyses yielded a decomposition into functional sub-networks:

VTA: The first component had an eigenvalue of 13.7, explaining 10.4% of variance. 28 principal components had eigenvalues above 1, explaining 74.6% of cumulative variance in VTA connectivity. Each of the first four components explained >5% of the whole variance and 31.3% of the variance in summation, and were thus chosen for regression analysis.

SNc: The first component had an eigenvalue of 15.4, explaining 11.7% of variance. 28 principal components had eigenvalues above 1, explaining 74.9% of cumulative variance in SNc connectivity. Each of the first four components explained >5%, and 32.0% in summation of the whole variance and were thus chosen for regression analysis.

Figure 3 gives a detailed overview on factor loadings by functional connections for each of the eight retained principal components/sub-networks. Components were labelled according to results obtained from Neuro-Query (see supplementary for details). For components VTA-PC3 and SNc-PC2, loadings roughly corresponded to mean functional connectivity, suggesting that these components reflected functional connectivity strength in dopaminergic resting-state networks, rather than sub-networks, respectively.

**Figure 3:**
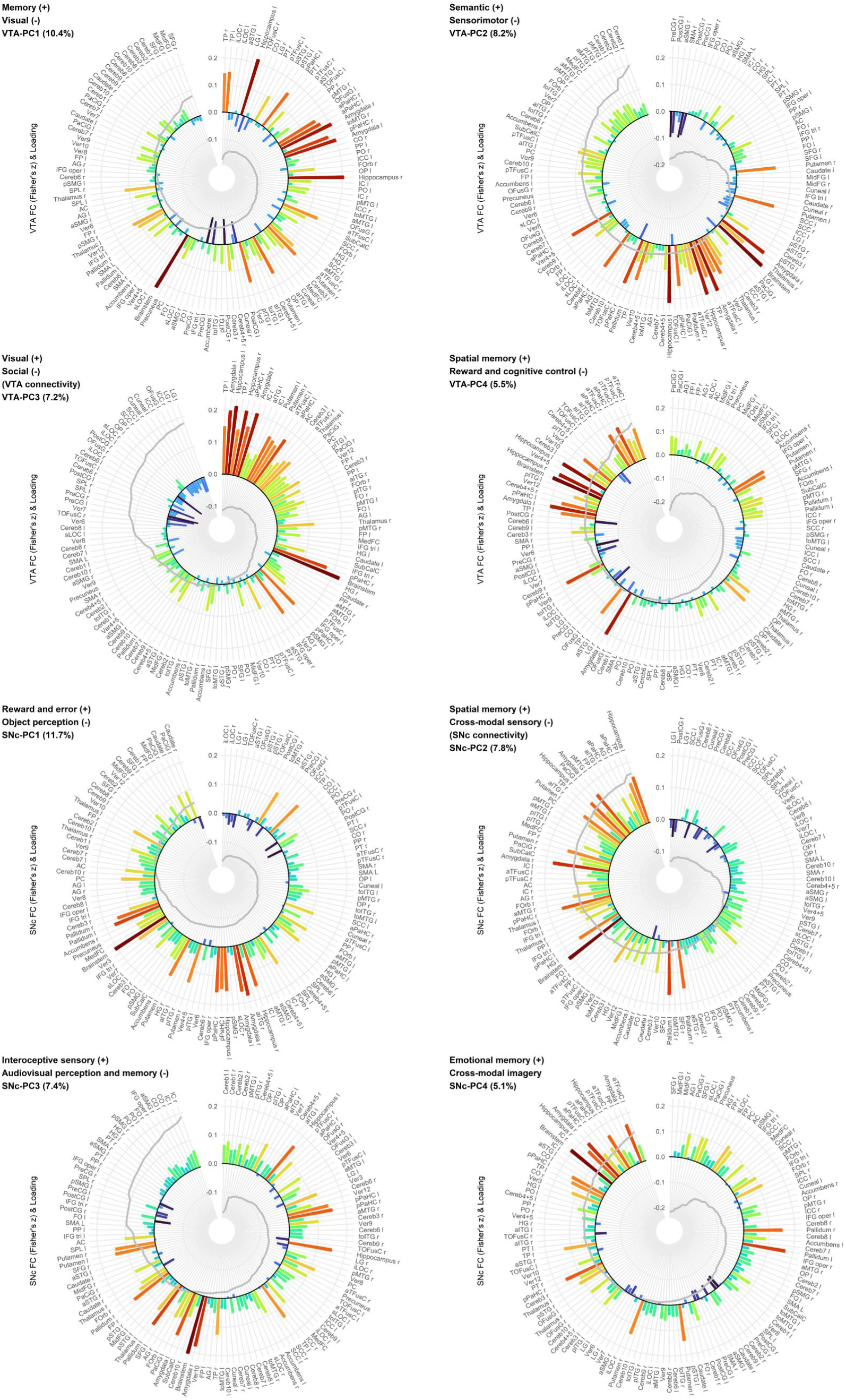
Radar charts describe the four components retained from PCA (sub-networks) of VTA (top row) and SNc (bottom row) functional connectivity by component loadings and preoperativ mean funcitonal connectivity. Loadings of functional connectivity (FC) between each seed ROI (VTA and SNc) to the respective target ROI (labels) on the components are superimposed as grey line graphs. Bar blots display sample mean functional connectivity (designated by height and colourization) between seed and target ROI. Sub-networks have been labelled according to a qualitative analysis with NeuroQuery. The label designated with (+) refers to functions assigned to target ROIs with positive component loadings, and the label designated with (-) refers to functions assigned to target ROIs with negative component loadings. Values in brackets are the amount of explained variance for the specific component. See also supplementary figures S3.

### 3.3 Preoperative functional connectivity

#### 3.3.1 Principal component regression (sub-networks)

The fourth sub-network (spatial memory^(+)^ – reward and cognitive control^(-)^) was significantly associated with POCD. Patients with higher values on this component had a decreased risk for POCD (B=-0.276, 95% CI=[-0.554; -0.124], OR=0.759 [0.575; 0.883], p=0.042).

The first SNc subnetwork (reward and error^(+)^ – object perception^(-)^) was significantly associated with POCD. Patients with higher values on this component had an increased risk for POCD (B=0.165 [0.028; 0.389], OR=1.180 [1.028; 1.476], p=0.042)

Figure 4 displays the associations between principal component scores and POCD. Detailed results of the full regression models are given in the supplement (table S1).

**Figure 4:**
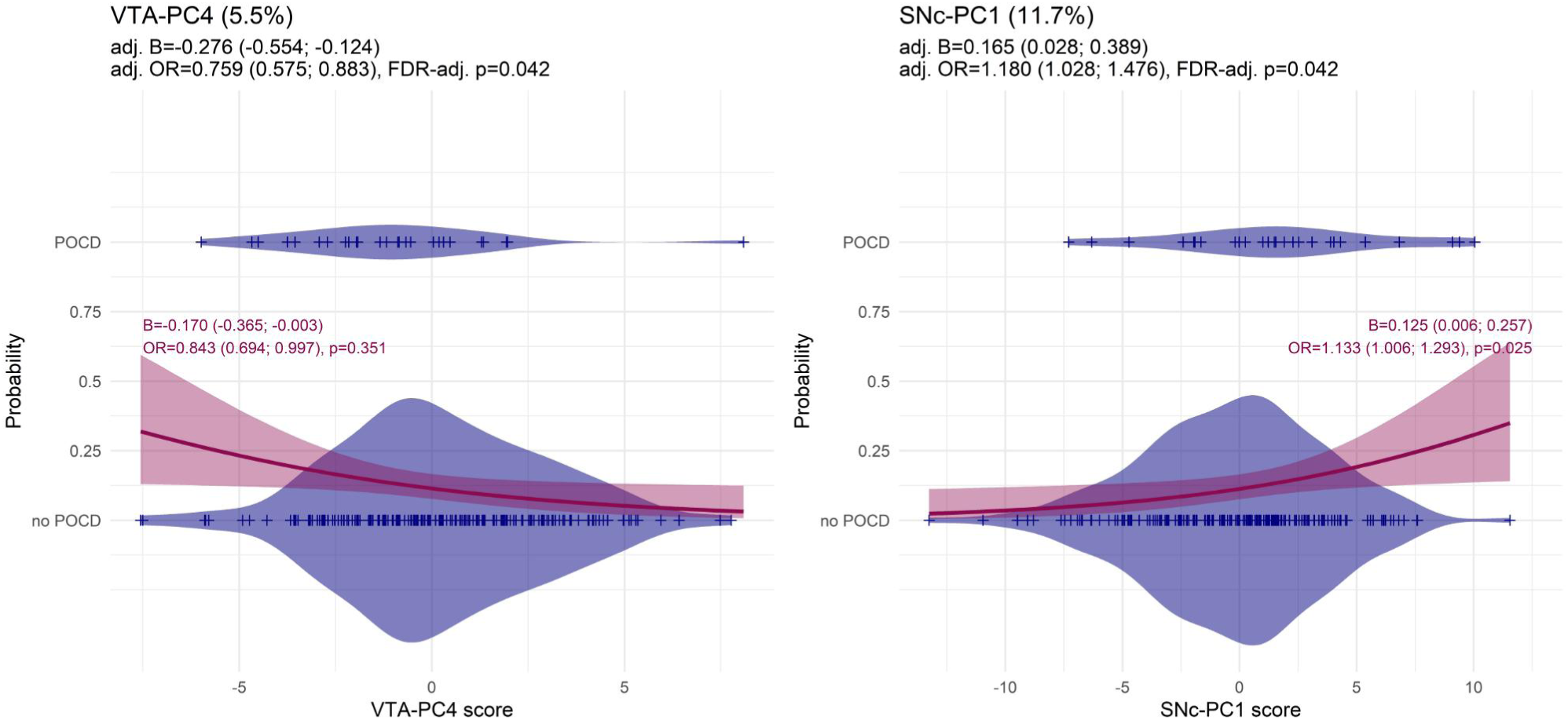
Association between scores on principal components displayed in figure 3 (i.e., PC4 for VTA functional connectivity, left, and PC1 for SNc functional connectivity, right, on the x-axes, respectively). Titles indicate the respective seed ROI and principal component with amount of explained variance in brackets. Violin and dot (+) plots display the observed distribution of principal component scores for patients with POD (upper violin plot) and patients without POD (lower violin plot). The red line displays the conditional probability of POCD (y-axis) depending on principal component score (x-axis), calculated from logistic regression with principal component score as the only independent variable. Red shaded areas indicate the 95% confidence interval. Results from principal component regression as described in the results section of the mansuscript are given below the title (regression coefficient B and odds ratio OR with 95% confidence intervals and p-value). In addition, the regression lines has been annotated with undadjusted results from logistic regression models with VTA-PC4 and SNc-PC1 as the only independent variables (red text, regression coefficient B and odds ratio OR with 95% confidence intervals and p-value). See also supplementary figure S4 for an ordinally scaled definition of POCD. Abbreviations: adj.; adjusted, FDR, false discovery rate

At an unadjusted level of significance, age and ASA physical status were positively associated with POCD occurrence. We conducted several exploratory analyses (supplementary material): We excluded four patients with intracranial surgery, which did not affect the findings substantially. We also repeated analyses stratified by sex .

#### 3.3.2 Voxelwise GLM

Voxelwise functional connectivity of the SNc did not differ between patients with POCD and control patients at baseline before surgery. For the VTA, one significant cluster of 118 voxels at (−06|-24|32) was observed indicating higher functional preoperative connectivity between the VTA and the cingulate gyrus (91 voxels in the posterior division and 14 voxels in the anterior division) in patients with POCD (supplementary figure S4). This cluster was not significant without additional adjustment for confounders.

### 3.4 Postoperative functional connectivity

#### 3.4.1 Principal component and multi-factor analysis (sub-networks)

After adjustment for multiple testing, none of the components of postoperative functional connectivity constructed from PCA and none of the factors derived from multi-factor analysis differed significantly between patients with POCD and control patients without POCD. Details are given in supplementary figure S5, tables S2 and S3.

#### 3.4.2 Voxelwise GLM

No differences in perioperative changes of VTA functional connectivity between patients with POCD and control patients have been observed. Functional connectivity of the SNc and a cluster at (−48|-34|12) covering 293 voxels in the left planum temporale, Heschl’s gyrus, the central and parietal operculum showed a stronger postoperative increase in patients with POCD compared to control patients both with and without adjustment for confounders (see supplementary figures S6 and S7).

## 4 Discussion

In this study, we analysed altered functional connectivity of the VTA and SNc in patients with POCD from large cohort undergoing elective surgery. Few studies have investigated VTA functional connectivity in POD (Choi *et al*. 2012; Oh *et al*. 2019), but studies for POCD were missing. We observed that both altered VTA and SNc functional connectivity in sub-networks predispose to POCD, but no functional connectivity alterations after surgery in patients with POCD.

### 4.1 VTA and SNc functional connectivity

VTA and SNc showed functional connectivity with vast cortical areas (supplementary figure S1). Although three major dopaminergic pathways originating from the mesencephalon are often stated in the literature (i.e., the mesocortical, the mesolimbic and the nigrostriatal systems), for both VTA (Berger, Gaspar and Verney 1991; Kwon and Jang 2014; Zubair *et al*. 2021) and SNc (Berger, Gaspar and Verney 1991; Kwon and Jang 2014; Zhang and Burock 2020) other vast and diffuse projections have also been described. Similar diffuse functional connectivity patterns had previously described in various populations by Giordano, Zhang and Peterson and a VTA-stimulation study in animals by Settell (Zhang *et al*. 2016; Peterson *et al*. 2017; Settell *et al*. 2017; Giordano *et al*. 2018). To the most extent, functional connectivity patterns in our study agree with previous reports. While functional imaging of brainstem regions is challenging, this notion is important, since the data presented by Giordano were collected at higher spatial resolution.

We used PCA and MFA to reduce the preoperative functional connectivity data and principal components may be interpreted as sub-networks of the VTA and SNc networks that are independent of each other and may show more or less prominent expression in individual patients.

### 4.2 Preoperative alterations in the dopaminergic sub-networks predispose to POCD

VTA and SNc preoperative functional sub-networks were associated with POCD. Altered functional connectivity of dopaminergic functional networks has already been described to be associated with cognitive function in older patients (De Marco and Venneri 2018), in Parkinson’s disease (Li *et al*. 2020; O’Shea *et al*. 2022; Zang *et al*. 2022) and in Alzheimer’s dementia (De Marco and Venneri 2018; Serra *et al*. 2018, 2021; Karim, Fahad and Rathore 2024). Our data suggest that preoperative dopaminergic dysfunction may contribute to cognitive decline after surgery and anaesthesia.

For both VTA and SNc, one principal component (i.e., sub-network) of preoperative functional connectivity was associated with POCD. Higher scores on the fourth principal component of VTA connectivity were protective against POCD, whereas higher scores on the first principal component of SNc connectivity increased the risk for POCD. The directions of principal components are arbitrary, but meaningful within the context of functional connectivity (i.e., correlated or anti-correlated BOLD signal fluctuations within an ROI) and factor loadings. From a simplified perspective, a target ROI with a high positive loading would contribute to a high score on the respective principal component when the BOLD signal fluctuations between seed and target ROI are strongly positively correlated (indicating simultaneous neuronal activation in seed and target ROI). Vice versa, a target ROI with a negative loading would also contribute to a high score on a principal component when the BOLD fluctuations between seed and target ROI are anticorrelated (indicating that the target ROI activation is particularly suppressed during seed ROI activation). Low scores on a principal component result from combinations of either negative loadings for target ROIs with correlated BOLD fluctuations or positive loadings for target ROIs with anticorrelated BOLD fluctuations.

Notably, we had denoted the respective components as related to *spatial memory^(+)^ - reward and cognitive control^(-)^* for the VTA and *reward and error^(+)^ - object perception^(-)^* for the SNc based on our analysis with NeuroQuery. In summary, higher dopaminergic connectivity to regions associated with spatial perceptive processes and lower connectivity to cognitive control-related areas may predispose to POCD.

With respect to the VTA, strong functional connectivity to the anterior and posterior fusiform cortex (aTFusC and pTFuSC), the anterior parahippocampal cortex (aPaHC) and anterior inferior temporal gyrus (aITG) seem to decrease risk for POCD. In contrast, strong connectivity of the VTA to the medial frontal cortex (MedFC), frontal orbital cortex (FOrb), frontal pole (FP), cingulate and paracingulate cortex (AC, PC and PaCig), angular gyrus (AG) and precuneus was associated with increased risk for POCD. In summary, higher preoperative VTA connectivity to temporal and hippocampal areas may protect against POCD, whereas strong integration of frontal, cingular and parietal areas into the VTA network would predispose for cognitive decline after surgery.

Areas with positive component loadings, i.e., the fusiform gyrus, the inferior temporal cortex and the parahippocampal gyrus, have been associated with spatial and source memory, as well as visual perception (Dichter *et al*. 2009; Dichter, Felder and Smoski 2010; Mai and Paxinos 2011; Aminoff, Kveraga and Bar 2013; Elward, Vilberg and Rugg 2015; Jääskeläinen *et al*. 2016; Landin-Romero *et al*. 2016; Pidgeon *et al*. 2016; Karanian and Slotnick 2017; Conway 2018; Burns, Arnold and Bukach 2019). Areas with negative component loadings are highly connected multimodal association areas often involved in salience and motivation and cognitive control, i.e., precuneus, medial frontal cortex (van den Heuvel and Sporns 2011), the angular gyrus, the precuneus and posterior cingulate cortex (Grabner *et al*. 2007, 2009; Bonner *et al*. 2013; Seghier 2013; Elward, Vilberg and Rugg 2015; Rolls 2019; Messina *et al*. 2023), the prefrontal and cingulate cortex (Dichter *et al*. 2009; Dichter, Felder and Smoski 2010; Rolls 2019; Friedman and Robbins 2022).

For the SNc, strong connectivity to the paracingulate cortex (PaCig) as well as frontal areas, and impaired suppression of caudate activity during SNc activity would contribute to high scores on the first component with increased risk for POCD, but most notably, a large cluster of brain regions with un- or anticorrelated BOLD fluctuations (i.e., neuronal suppression during SNc activation) had negative factor loadings. These regions are usually associated with motivation and reward monitoring, self reflection and recognition of intention (Bengtsson, Lau and Passingham 2009; Stanley, Gowen and Miall 2010; Schiffer and Schubotz 2011; Robinson *et al*. 2012; Tricomi and Fiez 2012; Li *et al*. 2013; Appelgren and Bengtsson 2015).

The cluster of supressed ROIs comprised the temporooccipital cortex (TOFusC), lingual gyrus (LG) and lower occipital cortex (iLOC), whereas the ROIs with more or less uncorrelated BOLD fluctuations cover the occipito-temporal cortex, i.e. the occipital fusiform gyrus (OFusG) and intracalcarine cortex (ICC) the lingual gyrus as well as parts of the anterior and posterior superior temporal gyrus (aSTG and pSTG) and temporooccipital middle temporal gyrus (toMTG). Strong neuronal suppression during SNc activity would contribute to a high score on the first principal component and hence increase the risk for POCD. The majority of these areas are involved in object perception (Hickok and Poeppel 2007; Nagy, Greenlee and Kovács 2012; Wu *et al*. 2012; Mizokami *et al*. 2014; Milner and Cavina-Pratesi 2018; Burns, Arnold and Bukach 2019).

Beyond cognitive functions, NeuroQuery suggested relevance of sub-networks for certain diseases, e.g., major depressive disorder (Dichter *et al*. 2009; Dichter, Felder and Smoski 2010; Ma *et al*. 2012; Watanabe *et al*. 2015). For SNc-PC1, NeuroQuery indicated also a relation to schizophrenia (Wysiadecki *et al*. 2021; Merola *et al*. 2024). However, we observed a similar pattern for the VTA-PC4: Especially the precuneus, orbital, cingulate and paracingulate cortex have been associated with schizophrenia and hallucinations (Wysiadecki *et al*. 2021; Messina *et al*. 2023; Merola *et al*. 2024). Patients with schizophrenia have increased dopaminergic neurotransmission during psychosis (McCutcheon *et al*. 2018), dopaminergic deficits may account for negative symptoms during remission (Avram *et al*. 2019; Brandl *et al*. 2023).

The association of POCD with dopaminergic connectivity to regions associated with schizophrenia and hallucinations is interesting, since POCD is more common in patients who have experienced POD (Norkienė *et al*. 2010; Franck *et al*. 2016), and dopaminergic dysfunction in these areas may equally account for symptoms in schizophrenia, POD and POCD. However, POD was rare in the investigated sample and POCD prevalence was not increased in patients with POD, questioning an overlap between these entities. Recent studies on VTA, and to a lesser extent, SNc connectivity in schizophrenia described predominantly reduced dopaminergic connectivity to subcortical and vast occipital, temporal, parietal, insular and frontal areas (Schulz *et al*. 2022), whereas in our data, only decreased functional connectivity between VTA and hippocampus, parahippocampal gyrus, postcentral cortex and fusiform cortex was associated with POCD.

The qualitative analysis with NeuroQuery indicated both function- and disease related expansion terms for target ROIs with extreme loadings on the VTA-PC4 and SNc-PC1. It has recently been pointed out that in primates, and especially in humans, the VTA and the SNc may not constitute distinct cell groups based on histological and functional studies (Düzel *et al*. 2009). Hence, we compared component loadings and scores of VTA-PC4 and SNc-PC1, respectively and found a good correlation, supporting the notion that these dopmanergic functional sub-networks are not independent (supplementary figure S8).

### 4.3 No evidence for postoperative alterations of dopaminergic connectivity in POCD

By using two different approaches to identify postoperative functional connectivity patterns, we identified no postoperative changes associated with POCD. Our results are limited as both approaches allow that counter-acting longitudinal changes in distinct functional connections cancel each other out. So far our results suggest that the devolopment of POCD may be related to a chronic condition which is present before surgery rather dopaminergic dysfunction induced by anaesthesia and surgery. This conclusion may further be supported by our notion that age, and ASA physical status as an indicator for the patient’s genereal preoperative health status were associated with POCD and that characteristics of surgery and anaesthesia had relatively little impact on POCD risk.

Previous studies on AD and amnestic MCI reported decreased functional connectivity of the VTA with the parietal lobe, precentral cortex, posterior cingulate cortex and precuneus (Serra *et al*. 2018, 2021). However, in our data, decreased functional connectivity of the posterior cingulate cortex, precuneus and sections of the parietal lobe (i.e., angular and supramarginal gyrus, superior parietal lobe, parietal operculum) were associated with decreased risk for POCD. In PD, decreased functional connectivity of the SNc with thalamus, pallidum, caudate nucleus and putamen were reported (Li *et al*. 2020; Zang *et al*. 2022). In our data, only decreased functional connectivity of the SNc with putamen and pallidum was associated with increased risk for POCD, whereas lower connectivity to the thalamus and caudate was found to be protective. These differences rather suggest that POCD is an entity distinct from AD or PD.

### 4.4 Comparison of sub-network analyses with voxelwise analyses

The rationale for analyses of sub-network analysis was founded on the assumption that dopaminergic nuclei have diffuse and rather sparse projections to vast cerebral target areas, and therefore, alterations in dopaminergic connectivity would be seen in a wide range of scattered target ROI, rather than in locally restricted clusters. However, as a cross check and for comparability with other studies, we conducted voxelwise analyses as well. We observed only spurious preoperative increases of VTA functional connectivity to the posterior and anterior cingulate cortex (AC, PC) in patients with POCD. Since AC and PC had high component loadings for VTA-PC4, this results provides little additional information, and rather suggests that the analysis of sub-networks is more efficient in this case.

While we observed no postoperative changes in preoperatively altered sub-networks related to POCD, voxel-wise analyses suggested and increase of postoperative functional connectivity between the SNc and a cluster covering left Heschl’s gyrus, planum temporale and the operculum. Notably, a meta-analysis reported progressive atrophy in the left Heschl’s gyrus and planum temporale in schizophrenia (Vita *et al*. 2012). While the type of antidopaminergic treatment with inhibition of neurotrophic factors has often been accused to modulate brain atrophy in schizophrenia (Lieberman *et al*. 2005; Vita *et al*. 2015; Hemby and McIntosh 2023). However, atrophy of the planum temporale in schizophrenia has also been found to correlate with duration of untreated psychosis (Keshavan *et al*. 1998; Takahashi *et al*. 2007). Dopaminergic-glutamatergic neurotoxicity has been suggested as a pathomechanism (Keshavan 1999). While it the reason for the particular vulnerability of the left superior temporal gyrus is unknown, dopaminergic-glutamatergic excitoxicity may also play a role for POCD. The identified cluster coincides with areas usually relevant for speech (Shapleske *et al*. 1999; Triarhou 2021; Henderson *et al*. 2023) and somatosensory areas (Eickhoff *et al*. 2006; Triarhou 2021). Due to its central function for understanding and production of speech and somatosenation, dysfunction of these arease may have detrimental impact on performance cognitive testing, including indirect effects on task understanding and sensory feedback during motoric task components.

Please note that, while we used widely-accepted standard levels of significance for Random-Field Theory in voxelwise analyses, these have not additionally been adjusted for analyses of two indenpendent seed ROIs.

### 4.5 Limitations

Functional MRI of infratentorial brain structures is challenging since brainstem nuclei are small and poorly contrasted in commonly used structural MRI sequences and artifacts from vessels nearby may arise. In the past, these problems have been adressed in methodologically outstanding studies using ultra-highfield MRI of the brain-stem providing sufficient contrast, image resolution below 1mm³ voxel size or cardiac-gated fMRI (D’Ardenne *et al*. 2008; Bianciardi *et al*. 2016; Hansen *et al*. 2024). Since these approaches are often associated with long acquisition times, additional risks and extraordinary discomfort for patients, they are often infeasible in surgical cohorts. Studies with more conventional functional imaging approaches describe a large variety of acquisition and analysis procedures, lacking a clear methodological gold standard. E.g., functional connectivity studies of the SN or VTA have used fMRI acquisition schemes with voxel sizes ranging between 13mm³ and 80mm³ (De Marco and Venneri 2018; Giordano *et al*. 2018; O’Shea *et al*. 2022). The majority of studies used probabilistic MRI atlases to define the ROIs, but only a minority reported the covered volumes. This is an important limitation in other studies, since ROI size may affect the results (Giordano *et al*. 2018). The smallest reported mask volume was 268mm³ for a ROI mask of the SNc (Karim, Fahad and Rathore 2024), and the largest reported volume was 1106mm³ for a combined mask of VTA and SNc (Zhang *et al*. 2016). Hence, most probabilistic masks, including the one used in this study, are significantly larger than the actual volume of the dopaminergic system. E.g., the average volume of the VTA has been reported as 60mm³ (D’Ardenne *et al*. 2008; Mai and Paxinos 2011). Studies using geometrical (i.e., spherical or cubic) masks to define dopaminergic seed ROIs often cover much smaller volumes (i.e., 34-113mm³ for the VTA), but all define the seed region at fixed coordinates in normalized images (Choi *et al*. 2012; De Marco and Venneri 2018; Giordano *et al*. 2018; Oh *et al*. 2019). Only one of this studies reports manual correction of ROIs in the brainstem, but not the VTA in particular (Oh *et al*. 2019). While this approach intends to reduce signal contamination from adjacent, non-dopaminergic regions, it bears the risk that the actual ROI is not covered, especially in the normalized images. Due to the lack of clear anatomical borders of the VTA and SNc in (normalized and smoothed) T1w images, we expect little benefit of manual correction of the seed ROI location, and the choice of ROI size reflects a specificity-sensitivity trade-off. While the inherent limitations of the approach have been outlined here, our study complies with the practiced methodological standard.

We here describe the functional connectivity patterns of the VTA and SNc to address dopaminergic dysfunction in POCD. However, the specificity of our approach for dopaminergic system alterations is limited, for several reasons: First, both VTA and SNc contain a substantial part of non-dopaminergic neurons (Mai and Paxinos 2011). Second, it is unknown if coactivation between the dopaminergic seed ROI and non-dopaminergic target ROI originates from the seed ROI and reflects dopamine release or originates from the target ROI and may hence be non-dopaminergic. Third, we do not know about non-dopaminergic relay structures. Functional connectivity between SNc and cortex may be indirect and mediated via the basal ganglia. Furthermore, dopamine is a vasoactive substance with direct effects on blood vessels, which may only partially be mediated by neuronal activity (Krimer *et al*. 1998). Finally, VTA and SNc are only a part of the brain’s dopaminergic system: The retrorubral fields (RRF, A8) also contain dopaminergic cells, and further minor groups of dopaminergic neurons are found in the periaqueductal grey (A11), zona incerta (A13), hypothalamus (A12, A14, A15) and the olfactory bulb (A16) as well as the retina (A17) (Mai and Paxinos 2011). For these, no probabilistic maps are available, and most of these regions are too small to provide robust signal in conventional fMRI.

Current guidelines for the assessment of postoperative neurocognitive disorders refer to either minor or major neurocognitive disorder (NCD) based on criteria from neuropsychological testing, impairment in daily activities and subjective cognitive concern (Evered *et al*. 2018)ast studies have assessed POCD based solely on cognitive trajectories (Abildstrom *et al*. 2000; Rasmussen *et al*. 2001; Borchers *et al*. 2021) and POCD assessment according to ISPOCD criteria was the scientific standard at the time of study conduction, but only considered neuropsychological testing as a sole criterion for POCD.

Principal component regression was used for data reduction and construction of sub-networks. The selected number of components retained for analysis was arbitrary, and explained only one-third of variance in VTA and SNc functional connectivity in total (5-10% per component). While this, on the one hand, may suggest that major artifacts and non-neuronal sources of fMRI signal fluctuation have been removed during preprocessing, it also means that most dopaminergic functional connectivity has been ignored in our analysis.

### 4.6 Conclusion

Altered preoperative functional connectivity of the dopaminergic system predisposes to POCD. Particularly relevant regions are associated with sensory, especially spatial perception, higher cognitive functions as well as regions thought to be involved in schizophrenia and hallucinations. We observed no associations of POCD with postoperative SNc or VTA functional connectivity, suggesting that dopaminergic network dysfunction is a predisposing, possibly chronic feature in POCD. However, VTA and SNc functional connectivity in patients with Alzheimer’s or Parkinson’s disease or schizophrenia shows little similarity with our results in POCD, suggesting that these conditions share little pathomechanistic features. To our knowledge, we here provide the first report on dopaminergic network dysfunction in POCD. Future studies may hence consider the dopaminergic system as a potential treatment and prevention target for POCD, but especially the question of dopaminergic hyper- or hypofunction in POCD remains further research. Studies on antidopaminergic treatment of POD will need to consider the potential longterm effects of their intervention on cognition as well.

## Data Availability

Participant data may be made available upon request following publication to researchers who provide a methodologically sound proposal in accordance with applicable legal and regulatory restrictions after careful review of each individual request. Access will only be granted in cases where the potential receiver of the data and purpose of the analysis is covered by the patients' informed consent and applicable legal regulations.
Proposals for data analysis must be directed to both. Analyses will be limited to those approved in appropriate ethics and governance arrangements. All study documents which do not identify individuals (e.g. study protocol, informed consent form template) will be freely available on request.

## Acknowledgements and disclosing remarks

We acknowledge the contributions of Pharmaimage Biomarker Solutions GmbH during the conduct of this study.

## 5 Statement on sex and gender

Sex, but not gender, was recorded as either male and female, without a standard operating procedure for the assessment of sex. Research staff was allowed to record sex according to both the patient’s self-report or the patient file. Analyses stratified for sex are given in the supplementary material.

## 6 Data availability statement

Participant data may be made available upon request following publication to researchers who provide a methodologically sound proposal in accordance with applicable legal and regulatory restrictions after careful review of each individual request. Access will only be granted in cases where the potential receiver of the data and purpose of the analysis is covered by the patients’ informed consent and applicable legal regulations.

Proposals for data analysis must be directed to both. Analyses will be limited to those approved in appropriate ethics and governance arrangements. All study documents which do not identify individuals (e.g. study protocol, informed consent form template) will be freely available on request.

A preprint of this article was posted to medrXiv (doi.org/10.1101/2025.01.02.25319918, on Dec 30 2024).

## 7 Author contributions statement

All authors contributed significantly to this study: Florian Lammers-Lietz: conceptualization, investigation, methodology, formal analysis, writing – original draft; Friedrich Borchers: methodology, investigation, formal analysis, data curation, validation; Insa Feinkohl: methodology, investigation, formal analysis; Cicek Kanar: formal analysis; Henning Krampe: investigation, writing – original draft; Gregor Lichtner: methodology; Claudia Spies: conceptualization, funding acquisition, data curation, resources, project administration; Jayanth Sreekanth: methodology; Janine Wiebach: methodology, formal analysis; Georg Winterer: conceptualization, funding acquisition, data curation, resources, project administration; Martin Weygandt: methodology; Friedemann Paul: conceptualization, supervision

## 10 Declaration of generative AI and AI-assisted technologies in the writing process

Artificial intelligence has not been used during the preparation of the manuscript. The supplementary section on fMRI preprocessing and functional connectivity analyses has been automatically been generated by CONN software and was edited by the authors.

## ABBREVIATIONS

AD: Alzheimer’s diseases
ASA(-PS): American Society of Anesthesiologists (physical status)
B: regression coefficient B
BioCog: Biomarker Development for Postoperative Cognitive Impairment in the Elderly study
BOLD: blood oxygenation level-dependent [signal]
CCI: Charlson’s comorbidity index
CI: confidence interval
EU: European Union
FC: functional connectivity
(f)MR(I): (functional) magnetic resonance (imaging)
GPT: grooved pegboard test
ISCED: international standard classification of education
ISPOCD: International Study of Post-Operative Cognitive Dysfunction
MFA: multi-factor analysis
MMSE: mini-mental status examination
NCD: neurocognitive disorder
OR: odds ratio
PAL: paired-associate learning test
PC(A): principal component (analysis)
PD: Parkinson’s disease
POCD: postoperative cognitive dysfunction
POD: postoperative delirium
RCI: reliable change index
ROI: region-of-interest
RRID: research resource identifier
RRF: retrorubral fields
sd: standard deviation
SNc: substantia nigra pars compacta
SRT: simple reaction test
SSP: simple span test
TMT(-A/B): trail making test (part A/B)
VRM: verbal recognition memory test
VTA: ventral tegmental area

A full list of abbreviations for brain regions is given in the supplementary material.

