## supplementary for "Dopaminergic sub-network connectivity alterations are associated with postoperative cognitive dysfunction: Results from the BioCog cohort study"

#### Abbreviations for brain regions

FP r, Frontal Pole Right

FP l, Frontal Pole Left

IC r, Insular Cortex Right

IC l, Insular Cortex Left

SFG r, Superior Frontal Gyrus Right

SFG l, Superior Frontal Gyrus Left

MidFG r, Middle Frontal Gyrus Right

MidFG l, Middle Frontal Gyrus Left

IFG tri r, Inferior Frontal Gyrus, pars triangularis Right

IFG tri l, Inferior Frontal Gyrus, pars triangularis Left

IFG oper r, Inferior Frontal Gyrus, pars opercularis Right

IFG oper l, Inferior Frontal Gyrus, pars opercularis Left

PreCG r, Precentral Gyrus Right

PreCG l, Precentral Gyrus Left

TP r, Temporal Pole Right

TP l, Temporal Pole Left

aSTG r, Superior Temporal Gyrus, anterior division Right

aSTG l, Superior Temporal Gyrus, anterior division Left

pSTG r, Superior Temporal Gyrus, posterior division Right

pSTG l, Superior Temporal Gyrus, posterior division Left

aMTG r, Middle Temporal Gyrus, anterior division Right

aMTG l, Middle Temporal Gyrus, anterior division Left

pMTG r, Middle Temporal Gyrus, posterior division Right

pMTG l, Middle Temporal Gyrus, posterior division Left

toMTG r, Middle Temporal Gyrus, temporooccipital part Right

toMTG l, Middle Temporal Gyrus, temporooccipital part Left

aITG r, Inferior Temporal Gyrus, anterior division Right

aITG l, Inferior Temporal Gyrus, anterior division Left

pITG r, Inferior Temporal Gyrus, posterior division Right

pITG l, Inferior Temporal Gyrus, posterior division Left

toITG r, Inferior Temporal Gyrus, temporooccipital part Right

toITG l, Inferior Temporal Gyrus, temporooccipital part Left

PostCG r, Postcentral Gyrus Right

PostCG l, Postcentral Gyrus Left

SPL r, Superior Parietal Lobule Right

SPL l, Superior Parietal Lobule Left

aSMG r, Supramarginal Gyrus, anterior division Right

aSMG l, Supramarginal Gyrus, anterior division Left

pSMG r, Supramarginal Gyrus, posterior division Right

pSMG l, Supramarginal Gyrus, posterior division Left

AG r, Angular Gyrus Right

AG l, Angular Gyrus Left

sLOC r, Lateral Occipital Cortex, superior division Right

sLOC l, Lateral Occipital Cortex, superior division Left

iLOC r, Lateral Occipital Cortex, inferior division Right

iLOC l, Lateral Occipital Cortex, inferior division Left

ICC r, Intracalcarine Cortex Right

ICC l, Intracalcarine Cortex Left

MedFC, Frontal Medial Cortex

SMA r, Supplementary Motor Cortex- Right

SMA L, Supplementary Motor Cortex- Left

SubCalC, Subcallosal Cortex

PaCiG r, Paracingulate Gyrus Right

PaCiG l, Paracingulate Gyrus Left

AC, Cingulate Gyrus, anterior division

PC, Cingulate Gyrus, posterior division

Cuneal r, Cuneal Cortex Right

Cuneal l, Cuneal Cortex Left

FOrb r, Frontal Orbital Cortex Right

FOrb l, Frontal Orbital Cortex Left

aPaHC r, Parahippocampal Gyrus, anterior division Right

aPaHC l, Parahippocampal Gyrus, anterior division Left

pPaHC r, Parahippocampal Gyrus, posterior division Right

pPaHC l, Parahippocampal Gyrus, posterior division Left

LG r, Lingual Gyrus Right

LG l, Lingual Gyrus Left

aTFusC r, Temporal Fusiform Cortex, anterior division Right

aTFusC l, Temporal Fusiform Cortex, anterior division Left

pTFusC r, Temporal Fusiform Cortex, posterior division Right

pTFusC l, Temporal Fusiform Cortex, posterior division Left

TOFusC r, Temporal Occipital Fusiform Cortex Right

TOFusC l, Temporal Occipital Fusiform Cortex Left

OFusG r, Occipital Fusiform Gyrus Right

OFusG l, Occipital Fusiform Gyrus Left

FO r, Frontal Operculum Cortex Right

FO l, Frontal Operculum Cortex Left

CO r, Central Opercular Cortex Right

CO l, Central Opercular Cortex Left

PO r, Parietal Operculum Cortex Right

PO l, Parietal Operculum Cortex Left

PP r, Planum Polare Right

PP l, Planum Polare Left

HG r, Heschl's Gyrus Right

HG l, Heschl's Gyrus Left

PT r, Planum Temporale Right

PT l, Planum Temporale Left

SCC r, Supracalcarine Cortex Right

SCC l, Supracalcarine Cortex Left

OP r, Occipital Pole Right

OP l, Occipital Pole Left

Cereb1 l, Cerebelum Crus1 Left

Cereb1 r, Cerebelum Crus1 Right

Cereb2 l, Cerebelum Crus2 Left

Cereb2 r, Cerebelum Crus2 Right

Cereb3 l, Cerebelum 3 Left

Cereb3 r, Cerebelum 3 Right

Cereb45 l, Cerebelum 4 5 Left

Cereb45 r, Cerebelum 4 5 Right

Cereb6 l, Cerebelum 6 Left

Cereb6 r, Cerebelum 6 Right

Cereb7 l, Cerebelum 7b Left

Cereb7 r, Cerebelum 7b Right

Cereb8 l, Cerebelum 8 Left

Cereb8 r, Cerebelum 8 Right

Cereb9 l, Cerebelum 9 Left

Cereb9 r, Cerebelum 9 Right

Cereb10 l, Cerebelum 10 Left

Cereb10 r, Cerebelum 10 Right

Ver12, Vermis 1 and 2

Ver3, Vermis 3

Ver45, Vermis 4 and 5

Ver6, Vermis 6

Ver7, Vermis 7

Ver8, Vermis 8

Ver9, Vermis 9

Ver10, Vermis 10

#### Supplementary methods: Neurocognitive testing

In total, the BioCog dataset provides data about POCD incidence after three months for 427 patients of the Berlin cohort.

POCD incidence was determined in relation to a non-surgical control group: 114 participants without indication for surgery but otherwise identical in- and exclusion criteria were invited to perform consecutive neuropsychological testing at baseline and three months to adjust for natural learning effects in cognitive testing [1]. Non-surgical control participants were recruited from outpatient clinics, primary care, elderly homes and via calls at public talks [2].

Testing was performed by trained study assistants in accordance with a standard operating procedure which was consented to by two neuropsychologists. Two independent assessors checked the data on plausibility including the review of free-text entries of research team members. When data for a participant were incomplete, missing values were imputed. If the data were missing due to impairment of concentration or poor understanding of test instruction according to the administering researcher, missing data were replaced with the worst performance value of the entire patient group, assuming that missingness reflects inability to perform the test due to cognitive decline. When values were missing at random, e.g. due to technical difficulties or environmental disturbances, random forest imputation was applied to replace missing values. Data were not imputed when neuropsychological testing was missing completely. The missForest package for R Statistical Software (RRID:SCR_001905) was used for imputations [3].

The following cognitive test parameters have been used for the calculation of POCD: mean correct latency from the Simple Reaction Time (SRT, processing speed), number of correctly remembered items in the free recall task (VRM free recall) as well as the number of correctly recognised items after delay on the Verbal Recognition Memory test (VRM recognition, verbal memory), span length in the Spatial Span task (SSP, working memory), the first trial memory score from the Paired Associate Learning test (PAL, visual memory) as implemented in the CANTAB test battery

(RRID:SCR_003001), as well as completion time of the Trail-Making-Test-B (TMT-B, executive functions) and completion time for the Grooved Pegboard test (GPT, fine motor skills). We selected these tests due to their moderate-to-good retest-reliability in the control group (intraclass coefficient between baseline and three months ≥0.75 based on a mean of multiple measurements, absolute-agreement, 2-way mixed-effects models) for this purpose [2]. Prior to calculation, SRT, GPT and TMT-B were log-transformed and sign-reversed to achieve an approximate normal distribution and a correspondence of higher scores with better cognitive performance. Details on single cognitive tests are given elsewhere [4].

For each cognitive test parameter, the corresponding RCI was calculated as $RCI=(\Delta X-\Delta X_c)/sd_{\Delta Xc}$. $\Delta X$ refers to the difference in test scores after surgery compared to baseline and $\Delta X_c$ refers to the mean test score difference between the corresponding measurement time points in the non-surgical control group. RCI is normalized to the standard deviation (sd) of mean differences in the control group $SD_{\Delta Xc}$. The compound RCI for each patient was defined as the sum of all RCIs in relation to the standard deviation of the sum of RCIs in the control group ($RCI_c$): compound $RCI=\Sigma(RCI)/sd_{\Sigma(RCIc)}$.

### Supplementary methods: Neuroimaging

#### Seed ROI definition

Substantia nigra, pars compacta (SNc, A9), the ventral tegmental area (VTA, A10) and the retrorubral fields (A8) are known to contain 76-90% of cerebral dopaminergic neurons (Mai and Paxinos 2011). Seed ROIs were defined by masks from the Brainstem Navigator 0.9 (Brainstem Imaging Lab, Athinoula A. Martinos Center for Biomedical Imaging) (Brainstem Navigator 2021), which had been created by semi-automatic and manual segmentations of multi-contrast MRI of living adult humans at 7 Tesla. Probabilistic atlases had been thresholded at 0.35 probability to create binary ROI masks (Bianciardi *et al.* 2015, 2018; García-Gomar *et al.* 2019, 2022; Singh *et al.* 2019; Singh, García-Gomar and Bianciardi 2021). The VTA has been defined according to the VTA_PBP_l/_r label (Ventral tegmental area – parabrachial pigmented nucleus complex) and the SNc has been defined according to the SN2_l/_r (Substantia nigra, subregion 2, compatible with compacta). We identified no ROI corresponding to the retrorubral fields, and hence, we ignored this large population of dopaminergic neurons (Mai and Paxinos 2011).

#### Functional MRI preprocessing and connectivity analyses

##### Preprocessing

Functional and anatomical data were preprocessed using a flexible preprocessing pipeline (Nieto-Castanon and Whitfield-Gabrieli) including realignment with correction of susceptibility distortion interactions, slice timing correction, outlier detection, direct segmentation and MNI-space normalization, and smoothing. Functional data were realigned using the SPM realign & unwarp procedure (Andersson *et al.* 2001), where all scans were coregistered to a reference image (first scan of the first session) using a least squares approach and a 6 parameter (rigid body) transformation (Friston *et al.* 1995), and resampled using b-spline interpolation to correct for motion and magnetic susceptibility interactions. Temporal misalignment between different slices of the functional data (acquired in descending order) was corrected following SPM slice-timing correction procedure (Henson *et al.* 1999; Sladky *et al.* 2011), using sinc temporal interpolation to resample each slice blood oxygenation level dependent signal (BOLD) time series to a common mid-acquisition time. Potential outlier scans were identified using ART (Whitfield-Gabrieli, Nieto-Castanon and Ghosh 2011) as acquisitions with framewise displacement above 0.5 mm or global BOLD signal changes above 3 standard deviations (Power *et al.* 2014; Nieto-Castanon 2022), and a reference BOLD image was computed for each subject by averaging all scans excluding outliers. Functional and anatomical data were normalized into standard MNI space, segmented into grey matter, white matter, and CSF tissue classes, and resampled to 3 mm isotropic voxels following a direct normalization procedure (Calhoun *et al.* 2017; Nieto-Castanon 2022) using SPM unified segmentation and normalization algorithm (Ashburner and Friston 2005; Ashburner 2007) with the default IXI-549 tissue probability map template. Last, functional data were smoothed using spatial convolution with a Gaussian kernel of 6 mm full-width half maximum (FWHM).

**Denoising**

In addition, functional data were denoised using a standard denoising pipeline (Nieto-Castanon and Whitfield-Gabrieli) including the regression of potential confounding effects characterized by white matter time series (5 CompCor noise components), CSF timeseries (5 CompCor noise components), motion parameters and their first order derivatives (12 factors) (Friston *et al.* 1996), outlier scans (below 237 factors) (Power *et al.* 2014), and linear trends (2 factors) within each functional run, followed by bandpass frequency filtering of the BOLD timeseries (Hallquist, Hwang and Luna 2013) between 0.008 Hz and 0.09 Hz. CompCor (Behzadi *et al.* 2007; Chai *et al.* 2012) noise components within white matter and CSF were estimated by computing the average BOLD signal as well as the largest principal components orthogonal to the BOLD average, motion parameters, and outlier scans within each subject's eroded segmentation masks. From the number of noise terms included in this denoising strategy, neuthe effective degrees of freedom of the BOLD signal after denoising were estimated to range from 8.5 to 140.4 (average 104.9) across all subjects (Nieto-Castanon 2022).

**Quality assessment**

Skull stripped T1 structural images were assessed by two independent raters (FLL and CK) according to a previously described protocol (Backhausen et al. 2016). Both researchers rated anatomical images on a scale (1=good, 2=moderate, 3=poor) concerning overall image sharpness, ringing artefacts, contrast to noise-ratio of the cortex and subcortical grey matter. Mean scores from both raters were averaged into a single score. For the quality assessment of functional images, we used indicators from another previously described protocol (Morfini, Whitfield-Gabrieli and Nieto-Castañón 2023). The assessment was modified to account for expected findings of brain atrophy in our sample: Rather than assessing grey and white matter volume as independent indicators for segmentation accuracy, we considered the ratio of grey to white matter as one aggregated indicator. Furthermore, the score from ratings of structural images was also integrated as an indicator of image quality into the assessment protocol. In accordance with the published protocol (Morfini, Whitfield-Gabrieli and Nieto-Castañón 2023), indicator values of >Q3+3·interquartile range (IQR) or <Q1-3·IQR were considered as outliers for indicators with unidirectional association with image quality, respectively, and indicator values outside the range of [Q1-1.5·IQR; Q3+1.5·IQR] were considered outliers for indicators with bidirectional association with image quality. Any patient with at least two outlying values was individually appraised for exclusion by one researcher (FLL).

### Supplementary results: Component interpretation with NeuroQuery

VTA-PC1

**NeuroQuery search:** *superior frontal gyrus, middle frontal gyrus, cerebellar cru*s (positive loadings, a)

**Expansion terms (similarity):** resting state (0.01), resting (0.01), working (0.01), verbal working (0.01), working memory (0.01), memory (0.01), dmn [default mode network] (0.01), task (0.01), force (<0.01), reward (<0.01), motion (<0.01), default (<0.01), finger (<0.01)

**NeuroQuery search:** *superior frontal gyrus, middle frontal gyrus* (positive loadings, b)

**Expansion terms (similarity):** resting state (0.02), resting (0.02), working memory (0.01), working (0.01), memory (0.01), task (0.01), DMN [default mode network] (0.01), reward (<0.01), default (<0.01), visual (<0.01), motor (<0.01), motion (<0.01), auditory (<0.01)

**Summary of publications related to the queries:** regions with extreme positive loadings were related to working memory (cerebellar crus[5–7,] , middle frontal gyrus[8]) and learnig/memory (cerebellar crus[9] and middle frontal gyrus[10,11]). Crus I and II are engaged in supramodal functional network including the prefrontal cortex[12,13].

**NeuroQuery search:** *temporal pole, lateral occipital cortex, superior temporal gyrus, fusiform cortex* (negative loadings)

**Expansion terms (similarity):** object (0.06), visual (0.06), action (0.05), face (0.04), semantic (0.04), empathy (0.04), social (0.03), verb (0.02), action observation (0.02), auditory (0.01), language (0.01), word (0.01), motor (0.01), hand (0.01), motion (<0.01)

**Summary of publications related to the query:** regions with extreme negative loadings were related to visual processing (lateral occipital cortex[14–17], fusiform cortex[16–21], superior temporal gyrus[20])

VTA-PC2

**NeuroQuery search:** *cerebellar crus, middle temporal gyrus, inferior temporal gyrus* (positive loadings)

**Expansion terms (similarity):** visual (0.01), resting state (0.01), semantic (0.01), resting (0.01), verbal working (0.01), chinese (0.01), object (<0.01), sentence (<0.01), DMN [default mode network] (<0.01), speech (<0.01), working (<0.01), face (<0.01), motion (<0.01)

**Summary of publications related to the query:** regions with extreme positive loadings were related to speech perception, reading and language processing (cerebellar crus[22–24], inferior[23] and middle temporal gyrus[22,23])

**NeuroQuery search:** *sensorimotor cortex, supplementary motor area, parietal operculum, supramarginal gyrus* (negative loadings)

**Expansion terms (similarity):** tactile (0.05), stimulation (0.03), dystonia (0.02), imitation (0.02), pain (0.02), phonological (0.02), movement (0.02), speech (0.02), language (0.02), sentence (0.01)

**Summary of publications related to the query:** regions with extreme negative loadings were related to sensory perception (postcentral gyrus[25–27] and parietal operculum[25–28]), and processing of sensory input (e.g., processing of phonological and haptic input in the supramarginal gyrus[26,29]) motor coordination (precentral gyrus[30] and supplementary motor area[30–32])

<u>VTA-PC3</u>

**NeuroQuery search:** *lingual gyrus, calcarine fissure, occipital pole* (positive loadings)

**Expansion terms (similarity):** visual (0.17), hallucination (0.04), acupuncture (0.03), stimuli (0.02), ASD [autism spectrum disorder] (0.02), gaze (0.02), saccade (0.01), stimulus (0.01), early visual (0.01), visual search (0.01), auditory hallucination (0.01), frequency (0.01), visual hallucination (0.01), MDD [major depressive disorder] (0.01), visual field (0.01), visual word (0.01), stimulation (<0.01), motion (<0.01), working (<0.01)

**Summary of publications related to the query:** regions with extreme positive loadings were related to visual perception and processing[33–35]

**NeuroQuery search:** *temporal pole, amygdala, hippocampus* (negative loadings)

**Expansion terms (similarity):** social (0.3), emotional (0.2), fear (0.2), theory [of] mind (0.2), TOM [theory of mind] (0.2), affective (0.01), empathy (0.01), mind (0.01), theory (0.01), game (0.01), sentence (0.01), neuroticism (0.01), fearful (0.01), migraine (0.01), sad (0.01), memory (<0.01)

**Summary of publications related to the query:** regions with extreme positive loadings were related to empathy (temporal pole[36,37]), emotional valence of cues and memories (amygdala[38–40], hippocampus[38–41]). Subregions of the temporal pole constitute functional networks with both hippocampus and the amygdala[42].

<u>VTA-PC4</u>

**NeuroQuery search:** *fusiform cortex, parahippocampal cortex, inferior temporal gyrus* (positive loadings)

**Expansion terms (similarity):** object (0.05), scene (0.04), spatial (0.04), memory (0.03), navigation (0.03), recollection (0.02), face (0.01), virtual (0.01), chinese (0.01), speech (<0.01)

**Summary of publications related to the query:** regions with extreme positive loadings were related to visual perception (inferior temporal gyrus[43], fusiform cortex[43,44], parahippocampal cortex[45]) and spatial or source memory (fusiform cortex[46], parahippocampal cortex[46,47])

**NeuroQuery search:** *paracingulate gyrus, angular gyrus, frontal pole* (negative loadings)

**Expansion terms (similarity):** reward (0.07), MDD [major depressive disorder] (0.05), cognitive control (0.04), major depressive disorder (0.03), depression (0.02), cognitive (0.02), meta (0.02), resting state (0.02), resting (0.01), semantic (0.01), sad (0.01), problem (0.01), reasoning (0.01), control (0.01), memory (0.01), task (0.01), default (<0.01), default mode (<0.01), DMN [default mode network] (<0.01), speech (<0.01)

**Summary of publications related to the query**: regions with extreme negative loadings were associated with cognitive control (frontal pole[48] and paracingulate gyrus[48]), reward feedback (angular gyrus[49], frontal pole[50] and paracingulate cortex[50]), emotional memory encoding and retrieval (frontal pole[51,52]) and higher-order cognitive functions (angular gyrus[53–55] and frontal pole[56])

SNc-PC1

**NeuroQuery search:** *caudate, paracingulate, middle frontal gyrus, superior frontal gyrus* (positive loadings)

**Expansion terms (similarity):** schizophrenia (0.04), resting state (0.04), resting (0.03), MDD [major depressive disorder] (0.03), self (0.2), theory mind (0.2), TOM [theory of mind] (0.2), reward (0.2), state (0.2), major depressive disorder (0.2), gaze (0.2), mind (0.2), familiar (0.2), theory (0.1), error (0.1), working (0.1), memory (0.1), task (0.1), movement (0.1), motion (0.1), DMN [default mode network] (0.1), default mode (0.1), default (0.1)

**Summary of publications related to the query:** regions with extreme positive loadings were related to reward/error monitoring and uncertainty (paracingulate gyrus, middle and superior frontal gyrus[57,] caudate[58–60]), self-reflection, motivation (paracingulate gyrus, middle and superior frontal gyrus[57]), and intention (paracingulate gyrus[57,59,61]). Connectivity analyses of the caudate nucleus suggested a cognition-related network including the middle frontal gyrus and a perception-related network including the superior frontal gyrus[62,63].

**NeuroQuery search:** *lateral occipital cortex, lingual gyrus, fusiform gyrus* (negative loadings)

**Expansion terms (similarity):** object (0.08), MDD [major depressive disorder] (0.01), perception (0.01), scene (0.01), default (<0.01), motion (<0.01)

**Summary of publications related to the query:** regions with extreme negative loadings were associated with visual perception (lingual gyrus[64], lateral occipital cortex[16,17] and fusiform gyrus[16,17])

SNc-PC2

**NeuroQuery search:** *hippocampus, temporal lobe, parahippocampal cortex* (positive loadings)

**Expansion terms (similarity):** scene (0.04), spatial (0.04), object (0.04), memory (0.03), navigation (0.03), epilepsy (0.03), temporal lobe epilepsy (0.02), recollection (0.01), virtual (0.01), egocentric (0.01), spatial navigation (0.01), location (0.01), item (0.01), spatial memory (0.01), AD [Alzheimer's disease] (0.01), Alzheimer (0.01), context (0.01), Alzheimer disease (0.01), semantic (0.01), speech (<0.01), default mode (<0.01), task (<0.01), default (<0.01), sound (<0.01), white (<0.01), voice (<0.01)

**Summary of publications related to the query:** regions with extreme positive loadings were related to memory (hippocampus)[65], including recognition of familiar items (hippocampus[66], inferior temporal cortex[66], putamen[66], temporal pole[66], posterior cingulate[66] and fusiform cortex[66]), but also especially spatial and contextual memory and navigation (hippocampus[67], parahippocampal cortex[45,46,67]), but also working memory for social cues (hippocampus[68], amygdala[68])

**NeuroQuery search:** *lingual gyrus, precentral gyrus, postcentral gyrus, pericalcarine cortex* (negative loadings)

**Expansion terms (similarity):** PD [Parkinson's disease] (0.04), Parkinson (0.03), Parkinson disease (0.03), cross modal (0.03), modal (0.03), blind (0.02), adolescencts (0.02), tactile (0.02), patient (0.02), disease (0.02), sentence (0.02), sighted (0.02), movement (0.01), hand (<0.01), motion (<0.01)

**Summary of publications related to the query:** regions with extreme negative loadings were associated with multi- or cross modal task performance, including written language comprehension (lingual gyrus[69,70]), language production (network including lingual gyrus, occipital and cerebellar regions[71]), hand grip control (somatosensory cortex[72], cerebellum[72]), complex visual stimulus features (somatosensory cortex[73], lingual gyrus[64,73] and cuneus[64]), visual perceptual load (somatosensory cortex[74])

SNc-PC3

**NeuroQuery search:** *insula, operculum, supramarginal gyrus* (positive loadings)

**Expansion terms (similarity):** somatosensory (0.04), taste (0.04), speech (0.04), food (0.03), tactile (0.02), phonological (0.02), touch (0.02), interoceptive (0.02), pain (0.02), obesity (0.02), pleasant (0.01), gustatory (0.01), hunger (0.01), sentence (0.01), task (0.01), reward (0.01), movement (0.01), hand (<0.01), sound (<0.01)

**Summary of publications related to the query:** regions with extreme positive loadings were related to gustatory sensations (insula[75,76], operculum[76]), vestibular sensation (insula[77]), pain sensation (insula[77–79], operculum[77]), somatosensation (insula[28,77], parietal operculum[27,28,77]), interoception (insula[77,80,81]) and emotion (insula[75,80–84]) and cognition (insula[81,83]). The dorsal insula formed a functional network with primary and secondary somatosensory areas including the supramarginal gyrus[85,86].

**NeuroQuery search:** *cerebellar crus, temporal lobe, occipital pole* (negative loadings, a)

**Expansion terms (similarity):** hallucination (0.05), ASD [autism spectrum disorder] (0.02), epilepsy (0.02), gaze (0.02), eye (0.02), saccade (0.02), temporal lobe epilepsy (0.01), auditory hallucination (0.01), verbal (0.01), visual hallucination (0.01), autism spectrum disorder (0.01), verbal working (0.01), force (0.01), timing (0.01), speech (<0.01), working (<0.01), DMN [default mode network] (<0.01), motion (<0.01), default (<0.01)

**Summary of publications related to the query:** regions with extreme negative loadings were associated with navigation (cerebellum[7], hippocampus[7]), autobiographic memory (cerebellum[9], hippocampus[9]). Functional connectivity analyses reported cerebellar networks with temporo-occipital regions[87].

**NeuroQuery search:** *ventral visual stream, cerebellum* (negative loadings, b)

**Expansion terms (similarity):** object (0.04), face (0.03), grasping (0.01), speech (0.01), recognition (0.01), scene (0.01), human (0.01), object recognition (0.01), dissociation (0.01), process (0.01), repetition (0.01), PMD (0.01), sound (0.01), word (<0.01), hand (<0.01), pain (<0.01), task (<0.01), working (<0.01), voice (<0.01), motion (<0.01)

**Summary of publications related to the query:** regions with extreme negative loadings were associated with a reading learning task (cerebellum[88], inferior temporal and fusiform cortex[88]) and perception of impossible figures (inferior temporal and fusiform gyrus[16])

SNc-PC4

**NeuroQuery search:** *fusiform cortex, amygdala, hippocampus, parahippocampal cortex,* (positive loadings)

**Expansion terms (similarity):** PTSD [posttraumatic stress disorder] (0.08), scene (0.04), object (0.03), memory (0.03), spatial (0.03), encoding (0.03), navigation (0.02), mindfulness (0.02), posttraumatic (0.02), posttraumatic stress disorder (0.02), trauma (0.02), recollection (0.01), traumati (0.01)c, sad (0.01), item (0.01), virtual (0.01), disorder (0.01), face (0.01), emotional (0.01), post traumatic stress disorder (0.01), subsequent memory (0.01), egocentric (0.01), location (0.01), spatial navigation (0.01), PPA [primary progressive aphasia] (0.01), task (<0.01), word (<0.01), working (<0.01), finger (<0.01), hand (<0.01), white (<0.01), motion (<0.01)

**Summary of publications related to the query:** regions with extreme positive loadings were related to contextual, especially spatial memory and navigation (hippocampus[89], parahippocampal cortex[45,47,89]), recognition, especially recognition of of faces (hippocampus[68,89,90], parahippocampal cortex[89,90], fusiform cortex[89,91]), as well as the perception of faces, especially in the context of emotional and social cues (fusiform cortex[92], amygdala[68,92])

**NeuroQuery search:** *frontal cortex, cingulate cortex, occipital cortex* (negative loadings)

**Expansion terms (similarity):** cross modal (0.04), modal (0.04), blind (0.03), sighted (0.02), cross (0.02), blindness (0.02), pain (0.02), tactile (0.01), acupuncture (0.01), fear (0.01), task (0.01), motion (<0.01), default (<0.01)

**Summary of publications related to the query:** regions with extreme negative loadings were associated with imagery (superior frontal gyrus[93,94], medial frontal cortex[93] and visual cortex[93,94]), cognitive control (cingulate cortex[95] and frontal areas[95]) and error monitoring (frontal[96,97] and cingulate areas[96,97])

### Supplementary figures

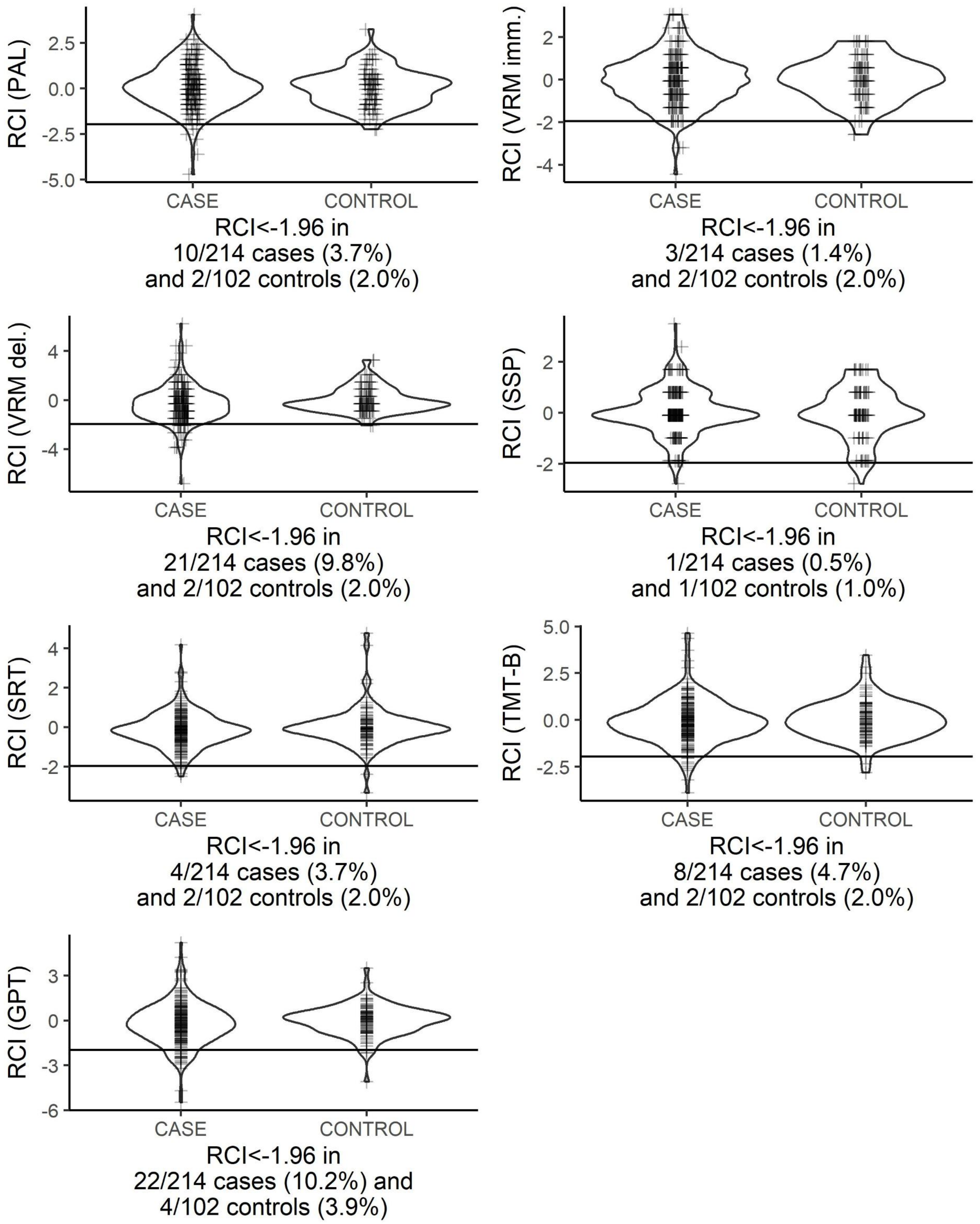

**Supplementary figure S1:** Violing plots and point plots of the reliable change index (RCI) after three months for all seven cognitive test parameters. The horizontal black line marks the used cut-off of -1.96. CASE here refers to 214 surgical patients undergoing anaesthesia, and CONTROL refers to participants without surgery. For better visualisation, data points of tests with discretely scaled outcomes (PAL, VRM, SSP) were jittered and bandwith for smoothing of the violin plot was increased by a factor of 1.5 for the SSP.

Abbreviations: PAL, Paired-Associate Learning; VRM, Verbal Recognition Memory; imm., immediate recall; del., delayed recognition; SSP, Simple Span Length; SRT, Simple Reaction Time; TMT-B, Trail-Making-Test pt. B; GPT, Grooved Pegboard Test

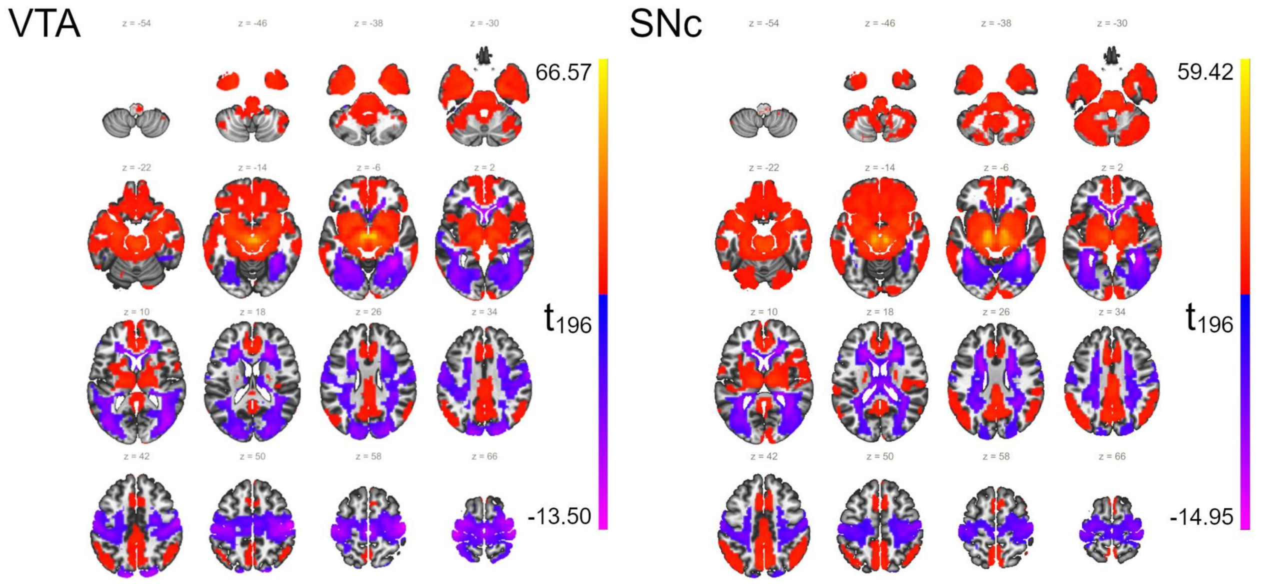

***Supplementary figure S2: Voxel-wise description of VTA (left) and SNc (right) functional networks in CONN. All voxels with significant functional connectivity to the seed ROI over all patients in the sample in a one-sample t-test are highlighted.** Please note that the maps describe VTA and SNc networks in the whole cohort rather than between group differences.*

*The figure displays voxel-wise t-values for difference of functional connectivity (Fisher's z) from 0, derived from a general linear model (GLM)[98].*

*Previous groups[99–101] describe a network comprising positive connections to prefrontal, pre- and postcentral, temporal, insular and (posterior) cingulate cortical areas, as well as the parahippocampal gyrus, thalamus, large portions of the basal ganglia, amygdala, the mesencephalon and cerebellum. Anticorrelated network parts in the occipital and posterior parietal cortex, middle and superior temporal gyri were also consistent between our work and the studies by Zhang and Peterson[99,100]. Compared to the study by Giordano, we found similar positive functional connections with the pallidum, ventral striatum, thalamus, midbrain and cerebellum, the cingulate cortex, ventrolateral prefrontal cortex, parahippocampal and inferior temporal gyri, the insula while there was discordance regarding pre- and postcentral areas[101]. The results by Giordano show substantial overlap with regions which respond to deep brain stimulation of the VTA in an animal experiment with both increase and suppression of the BOLD response, i.e., prefrontal cortex, limbic areas, primary sensorimotor areas, and the basal ganglia[102].*

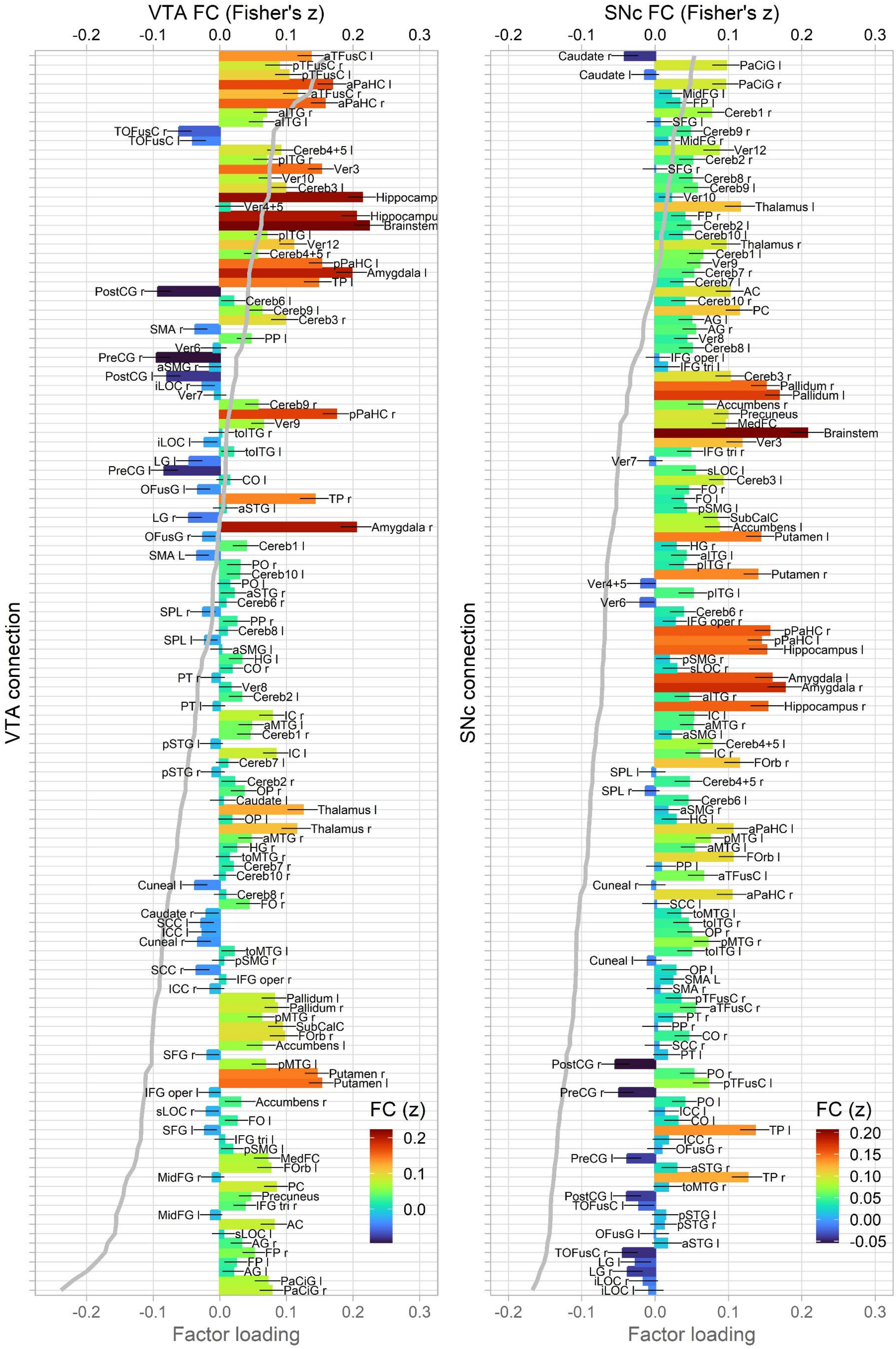
VTA FC (Fisher's z)
SNc FC (Fisher's z)
-0.2
-0.1
0.0
0.1
0.2
0.3
VTA connection
SNc connection
Factor loading
Factor loading
FC (z)
0.2
0.1
0.0
FC (z)
0.20
0.15
0.10
0.05
0.00
-0.05

**Supplementary figure S3:** Construction of principal components. PC4 for VTA (Spatial memory$^{(+)}$ – Reward and cognitive control$^{(-)}$, left) and PC1 for SNc (Reward and error$^{(+)}$ - object perception$^{(-)}$, right) were found to be significantly associated with POCD risk in logistic regression analyses. Loadings of functional connectivity (FC) between each seed ROI (VTA and SNc) to the respective target ROI (labels) on the components are superimposed as grey line graphs. Bar blots display sample mean functional connectivity (designated by height and colourization) between seed and target ROI. Black lines indicated 95% confidence intervall for mean FC. Connections are ordered by principal component loadings.

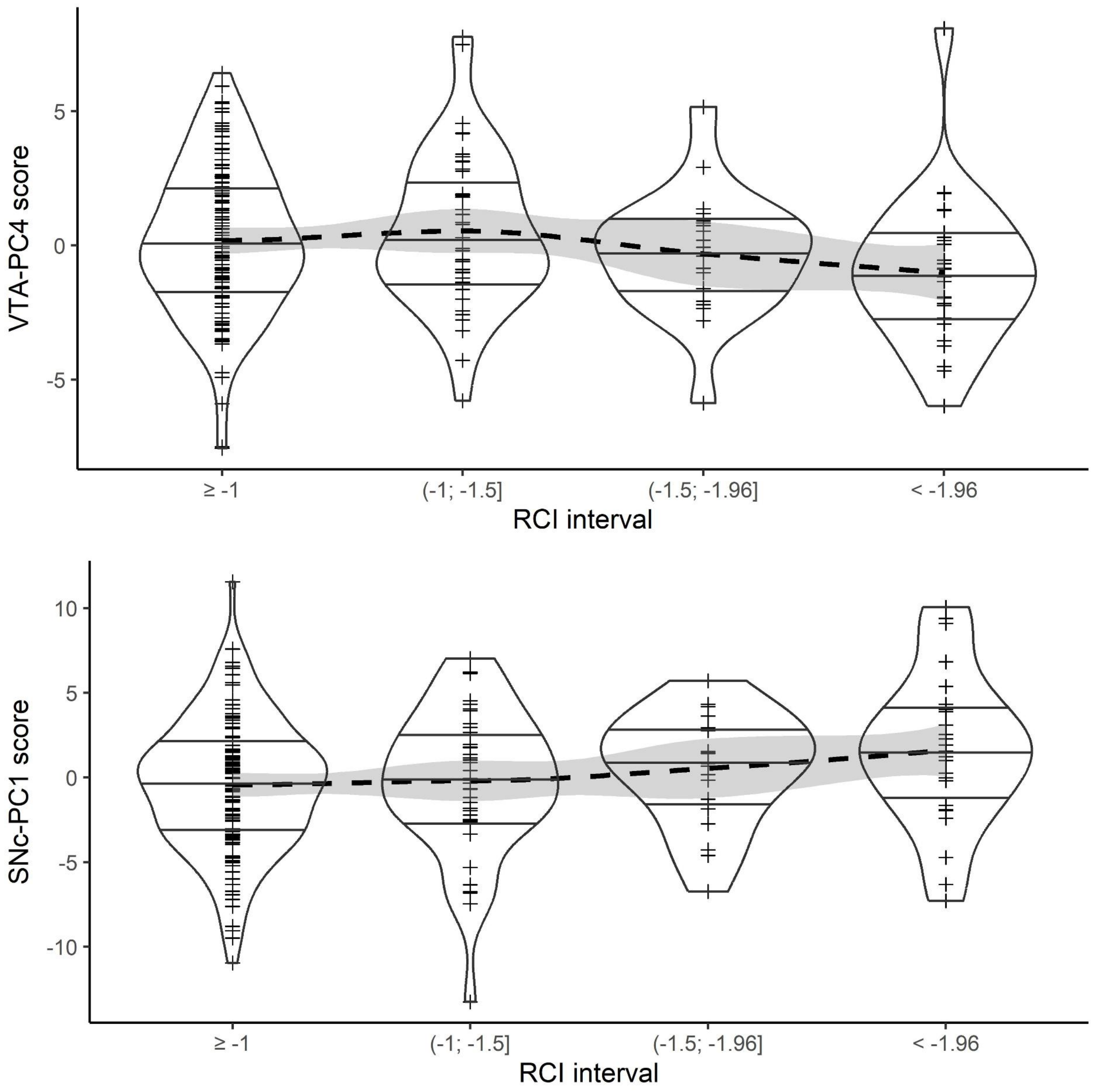

**Supplementary figure S4: Association between principal component scores as displayed in figure 4 and POCD at different cut-off values for the reliable change index (RCI).** For each interval, the component score is displayed as point and violin diagrams. For orientation, a LOESS (locally estimated scatterplot smoothing) regression graph is given as well (dashed line). Spearman's ρ is -0.11 (p=0.10) for VTA-PC4, and 0.15 (p=0.022) for the SNc-PC1.

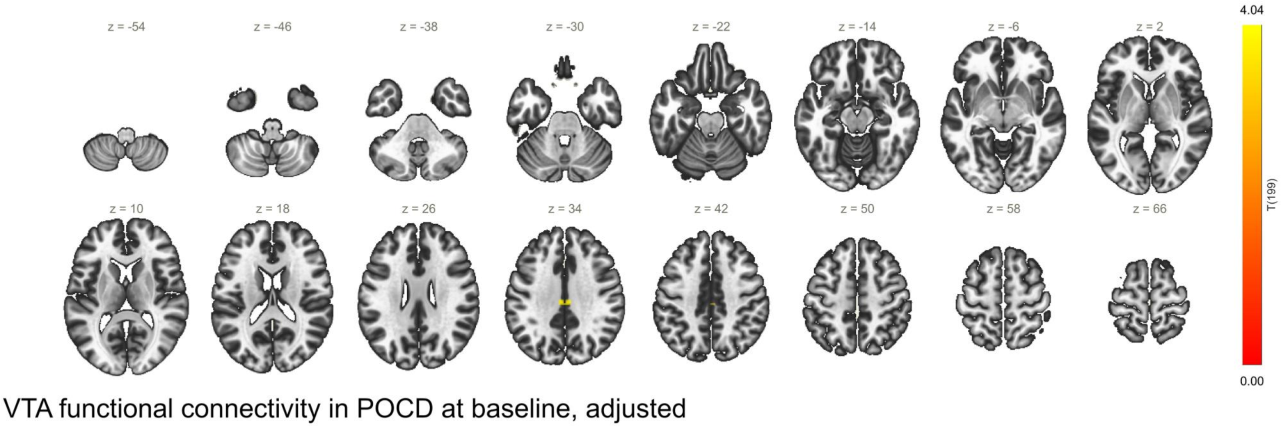

***Supplementary figure S4: Seed-based connectivty map of preoperative alterations between the VTA and the cingulate cortex in POCD.*** *Without adjustment for confounding variables, no significant clusters have been observed.*

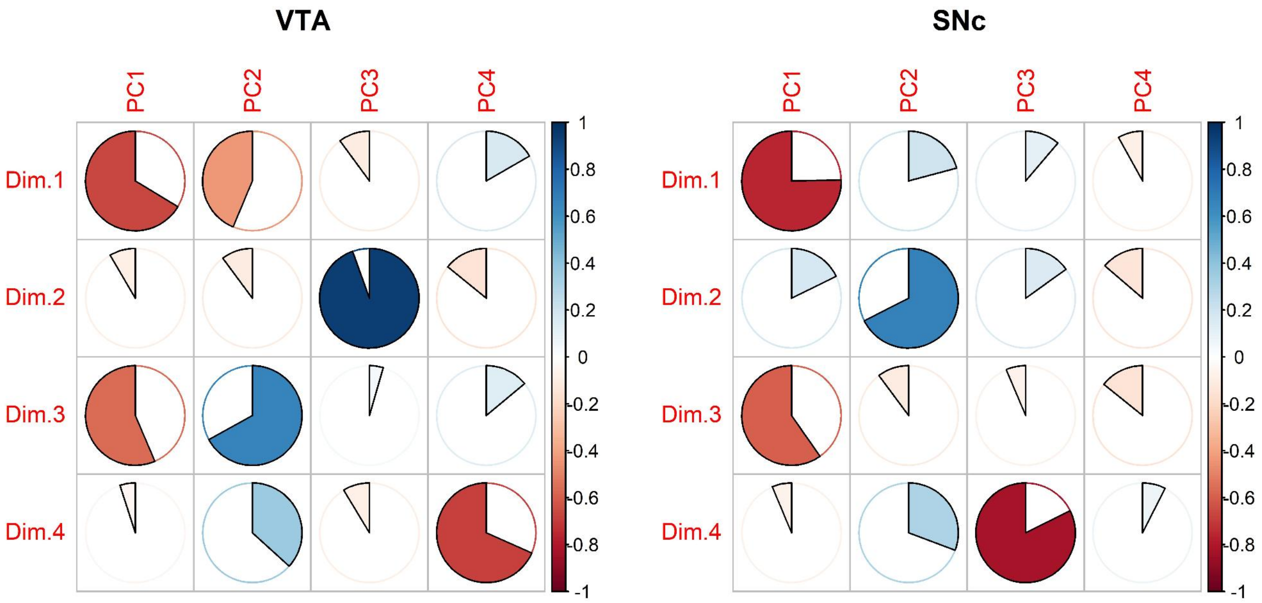

**Supplementary figure S5: Correlation matrix (Pearson's R) between scores on dimensions (i.e., factors) from MFA (multi-factor analysis) of longitudinal functional connectivity (Dim.1-Dim.4) and principal components of preoperative functional connectivity (PC1-PC4) for the ventral tegmental area (VTA, left) and the substantia nigra pars compacta (SNc, right).** Colours and pie charts indicate Pearson's R of the correlation. Please note that the orientation for components and factors in PCA and MFA is arbitrary.

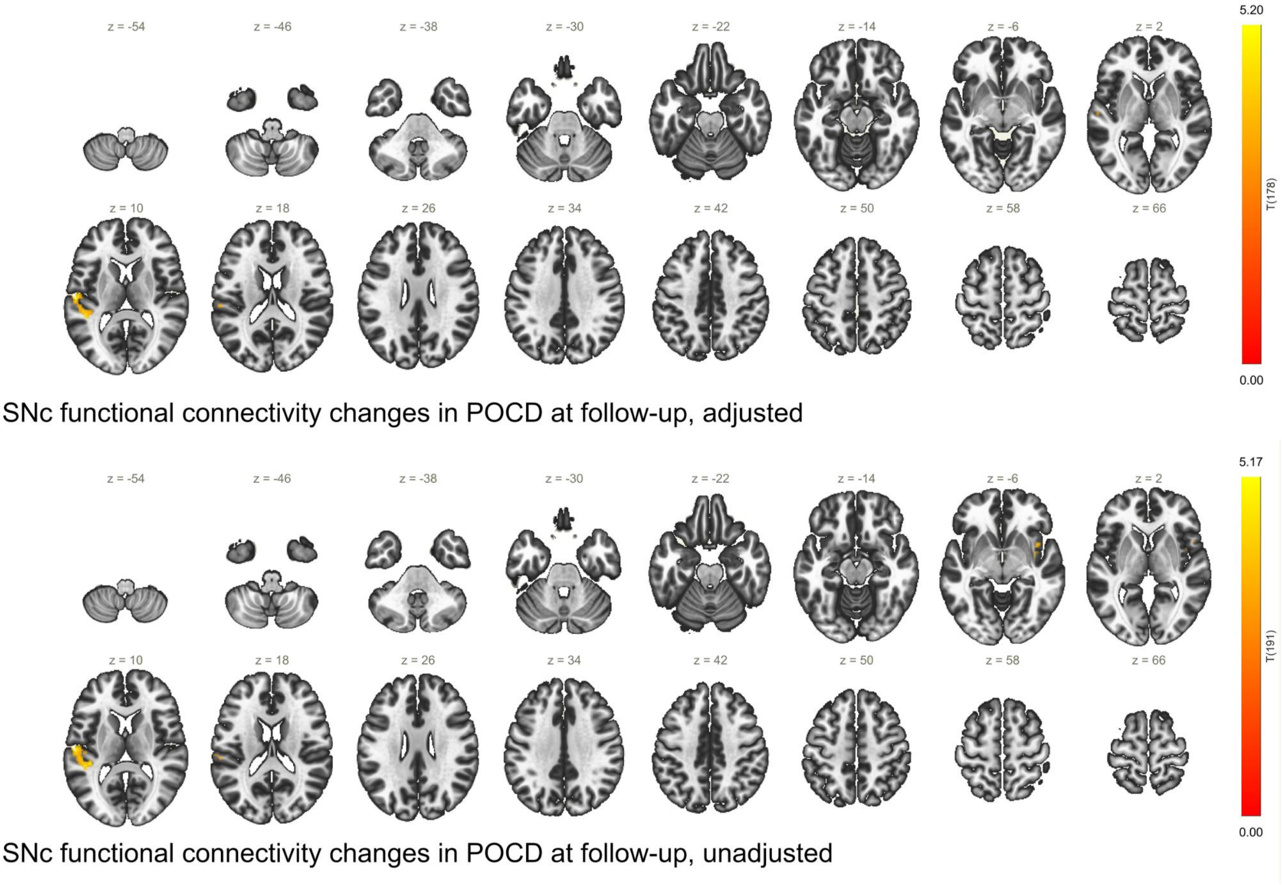

***Supplementary figure S6: Seed-based connectivty map of group by time interaction for the SNc. Functional connectivity between the SNc and the significant cluster in the left temporal lobe/operculum shows a stronger postoperative increase in POCD patients compared to control patients.*** *The upper panel provides GLM results after adjustment for confounders, whereas the lower panel has not been adjusted.*

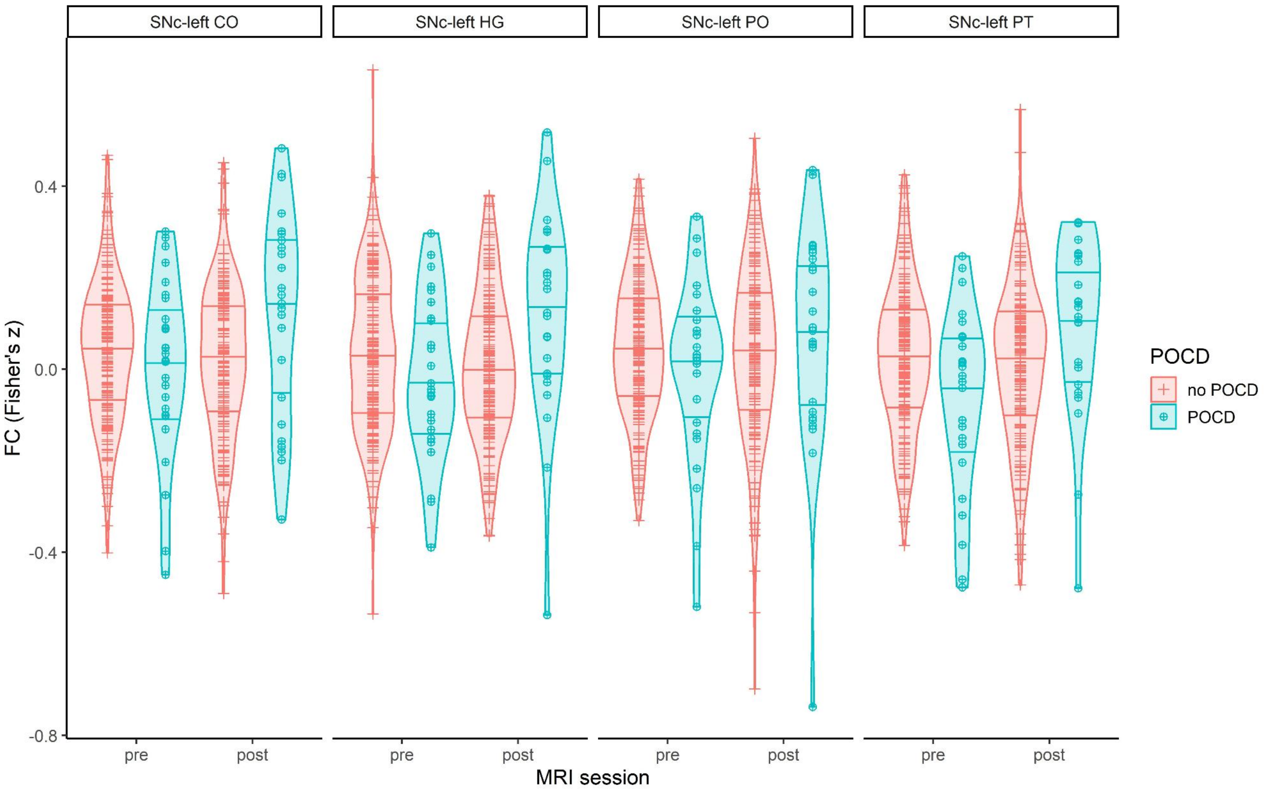

***Supplementary figure S7: Violin plots of seed-to-ROI functional connectivity between the SNc and four target ROIs covered by the cluster displayed in supplementary figure S6.*** *Control patients are displayed as orange crosses, patients with POCD are indicated in blue circled crosses. Horizontal lines mark the 25th, 50th and 75th percentiles.*

*Abbreviations: CO, central operculum; FC, functional connectivity; HG, Heschl's gyrus; PO, parietal operculum; PT, planum temporale*

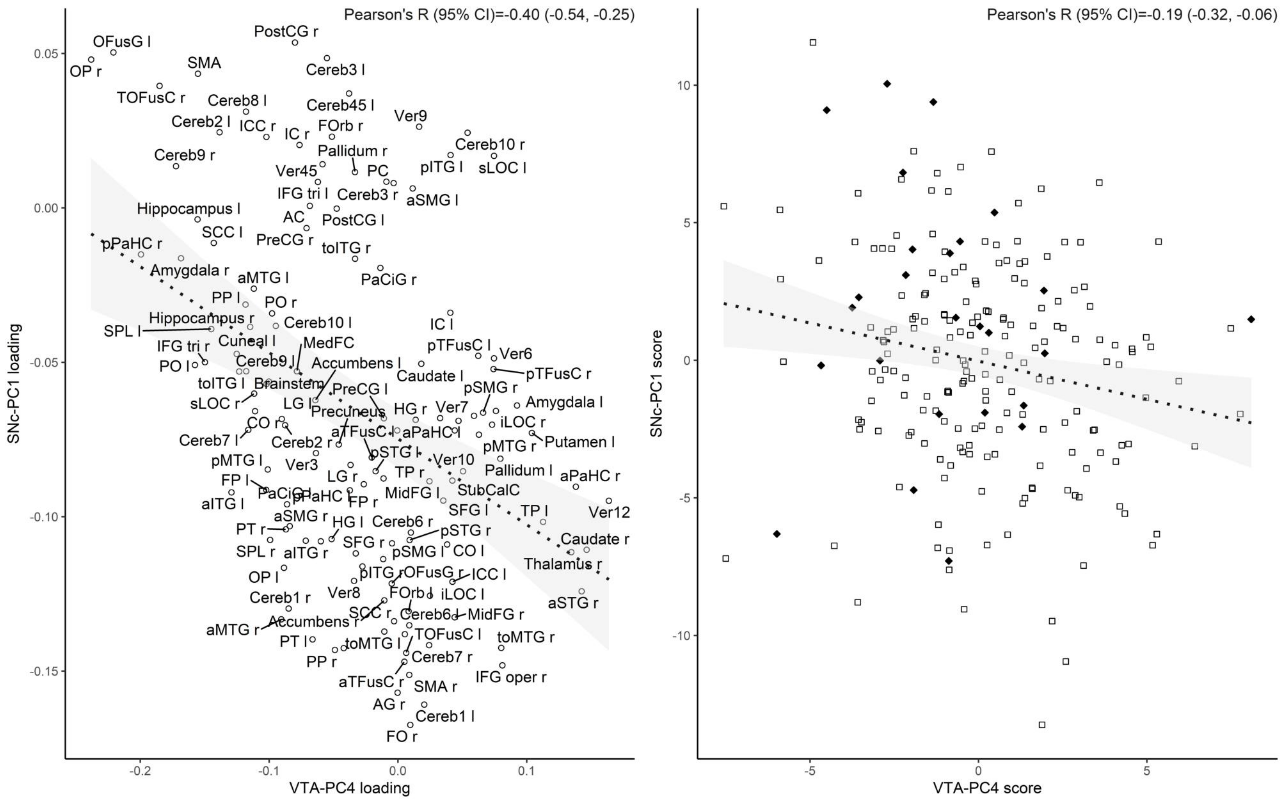

***Supplementary figure S8: Correlation of component loadings (left) and component scores (right) between VTA-PC4 (x-axis) and SNc (y-axis).*** *In the left figure, each point corresponds to a target ROI and the value on the x-/y-axes reflects the association of the seed ROI (SNc or VTA, respectively) with the respective principal component. In the right figure, each point corresponds to one patient. Patients with POCD are marked as ♦ and control patients as □. Patients with POCD have higher scores on SNc-PC1 and lower scores on VTA-PC4.*

*In both cases, significant correlations were observed. Pearson's R with 95% confidence interval is indicated in the top right corner of each figure. In conjunction with results from the qualitative functional analysis with NeuroQuery, these findings suggest that VTA-PC4 and SNc-PC1 reflect the same biological process.*

### Supplementary tables

**Supplementary table S1:** Associations of preoperative functional connectivity with POCD (N=214). Level of significance ($p<0.05$) was adjusted for eight independent tests (four principal components from two ROIs) using Benjamini-Hochberg correction ($p_{adj.}$) on 200/213 degrees of freedom. * indicates significance after adjustment for multiple testing.

| | **VTA** | | | **SNc** | | |
|---|---|---|---|---|---|---|
| | **B (95% CI)** | **OR (95% CI)** | **p ($p_{adj.}$)** | **B (95% CI)** | **OR (95% CI)** | **p ($p_{adj.}$)** |
| **PC1** | 0.052 (-0.141; 0.247) | 1.053 (0.869; 1.281) | 0.46 (0.8) | ***0.165 (0.028; 0.389)*** | ***1.180 (1.028; 1.476)*** | ***0.011 (0.042)**** |
| **PC2** | 0.046 (-0.167; 0.265) | 1.047 (0.846; 1.304) | 0.5 (0.8) | 0.022 (-0.152; 0.205) | 1.023 (0.859; 1.227) | 0.8 (0.8) |
| **PC3** | -0.022 (-0.185; 0.140) | 0.978 (0.831; 1.150) | 0.7 (0.8) | -0.032 (-0.248; 0.146) | 0.968 (0.780; 1.157) | 0.7 (0.8) |
| **PC4** | ***-0.276 (-0.554; -0.124)*** | ***0.759 (0.575; 0.883)*** | ***0.0057 (0.042)**** | -0.052 (-0.285; 0.179) | 0.950 (0.752; 1.197) | 0.6 (0.8) |
| **Age (years)** | 0.108 (0.009; 0.241) | 1.113 (1.009; 1.272) | 0.026 | 0.101 (-0.005; 0.251) | 1.106 (0.995; 1.285) | 0.038 |
| **Female sex** | -0.381 (-1.829; 0.769) | 0.683 (0.161; 2.158) | 0.44 | -0.355 (-1.767; 0.755) | 0.701 (0.171; 2.127) | 0.47 |
| **MMSE (points)** | -0.037 (-0.391; 0.503) | 1.038 (0.676; 1.653) | 0.8 | 0.108 (-0.358; 0.670) | 1.114 (0.699; 1.954) | 0.6 |
| **Duration of surgery (min)** | -0.004 (-0.015; 0.001) | 0.996 (0.986; 1.001) | 0.15 | -0.004 (-0.014; 0.000) | 0.996 (0.986; 1.000) | 0.13 |
| **ASA** | 1.256 (0.188; 2.746) | 3.511 (1.206; 1.557) | 0.012 | 1.252 (0.184; 2.719) | 3.499 (1.202; 15.167) | 0.013 |
| **Intracranial surgery** | 20.64 (17.297; 26.601) | $9\cdot10^{8}$ ($3.3\cdot10^{7}$; $3.6\cdot10^{11}$) | >0.9 | 17.45 (16.296; 22.625) | $3.8\cdot10^{7}$ ($1.2\cdot10^{7}$; $6.7\cdot10^{9}$) | >0.9 |
| **Intracavitary surgery** | 0.058 (-1.291; 1.289) | 1.059 (0.275; 0.363) | 0.9 | -0.006 (-1.330; 1.089) | 0.942 (0.264; 2.971) | 0.9 |
| **Regional anesthesia** | 0.073 (-0.759; 17.986) | 2.069 (0.468; $6.5\cdot10^{7}$) | 0.38 | 1.121 (-0.339; 18.650) | 3.069 (0.712; $1.3\cdot10^{8}$) | 0.18 |
| **Combined anesthesia** | 0.069 (-16.381; 18.577) | 2.000 ($7.7\cdot10^{-8}$; $1.2\cdot10^{8}$) | 0.6 | 0.944 (15.924; 18.947) | 2.571 ($1.2\cdot10^{-7}$; $1.7\cdot10^{8}$) | 0.49 |
| **Intercept** | -13.88 | | | -15.61 | | |

**Supplementary table S2:** Associations of POCD (N=193) with postoperative functional connectivity of the VTA and SNc. Postopertive components were calculated by multiplying component loadings from preoperative PCA with functional connectivity at follow-up after surgery. Level of significance ($p<0.05$) was adjusted for eight independent tests (four principal components from two ROIs) using Benjamini-Hochberg correction ($p_{adj.}$) on 180/192 degrees of freedom. B: linear regression coefficent, CI: confidence interval, POCD: postoperative cognitive dysfunction, PC: principal component, SNc: substantia nigra pars compacta, VTA: ventral tegmental area

| **Dependent variable** | **Independent variable** | **VTA** | | **SNc** | |
|---|---|---|---|---|---|
| | | **B (95% CI)** | **p ($p_{adj.}$)** | **B (95% CI)** | **p ($p_{adj.}$)** |
| **Postoperative component 1** | **POCD** | -0.023 (-0.357; 0.299) | 0.9 (>0.9) | -0.010 (-0.428; 0.421) | >0.9 (>0.9) |
| | **Preoperative PC score** | 0.044 (-0.001; 0.105) | 0.028 | 0.043 (0.016; 0.068) | 0.0034 |
| | **Interaction** | -0.013 (-0.103; 0.109) | 0.7 (0.9) | -0.005 (-0.118; 0.105) | 0.9 (0.9) |
| **Postoperative component 2** | **POCD** | -0.015 (-0.302; 0.268) | 0.9 (>0.9) | 0.172 (-0.071; 0.408) | 0.16 (0.6) |
| | **Preoperative PC score** | 0.038 (-0.014; 0.077) | 0.031 | 0.043 (0.018; 0.071) | 0.0005 |
| | **Interaction** | 0.039 (-0.066; 0.118) | 0.26 (0.9) | 0.021 (-0.052; 0.117) | 0.6 (0.9) |
| **Postoperative component 3** | **POCD** | -0.071 (-0.301; 0.162) | 0.5 (>0.9) | 0.212 (-0.056; 0.464) | 0.095 (0.6) |
| | **Preoperative PC score** | 0.038 (0.011; 0.093) | 0.027 | 0.040 (0.013; 0.067) | 0.0013 |
| | **Interaction** | 0.006 (-0.048; 0.118) | 0.9 (0.9) | -0.019 (-0.110; 0.074) | 0.7 (0.9) |
| **Postoperative component 4** | **POCD** | -0.027 (-0.242; 0.201) | 0.8 (>0.9) | 0.131 (-0.086; 0.356) | 0.24 (0.6) |
| | **Preoperative PC score** | 0.040 (0.000; 0.085) | 0.049 | 0.031 (0.003; 0.055) | 0.021 |
| | **Interaction** | -0.021 (-0.100; 0.069) | 0.6 (0.9) | -0.090 (-0.197; 0.035) | 0.044 (0.35) |

**Supplementary table S3:** Associations of POCD (N=193). Factor scores were derived from multi-factor analysis of pre- and postoperative functional connectivity. Level of significance ($p<0.05$) was adjusted for eight independent tests (four factors from two ROIs) using Benjamini-Hochberg correction ($p_{adj.}$) on 182/192 degrees of freedom. B: linear regression coefficent for POCD, CI: confidence interval, SNc: substantia nigra pars compacta, VTA: ventral tegmental area

| **Dependent variable** | **VTA** | | **SNc** | |
|---|---|---|---|---|
| | **B (95% CI)** | **p ($p_{adj.}$)** | **B (95% CI)** | **p ($p_{adj.}$)** |
| **Factor 1** | -0.393 (-1.031; 0.213) | 0.16 (0.32) | -0.533 (-1.145; 0.071) | 0.048 (0.19) |
| **Factor 2** | -0.093 (-0.624; 0.398) | 0.7 (0.8) | 0.396 (-0.069; 0.902) | 0.078 (0.21) |
| **Factor 3** | -0.113 (-0.657; 0.389) | 0.6 (0.8) | -0.194 (-0.672; 0.284) | 0.38 (0.6) |
| **Factor 4** | 0.558 (0.114; 1.031) | 0.011 (0.8) | -0.059 (-0.485; 0.359) | 0.8 (0.8) |

### Sensitivity analyses

#### Rationale and approach

Since we noticed the extreme regression coefficient and associated odds' ratio for intracranial surgery, we conducted sensitivity analyses to evaluate overfitting and influential outliers. The extreme estimates were caused by the fact that the only two patients with intracranial surgery in this sample developed POCD, whereas in the comparator group of patients without POCD, no patient had undergone intracranial surgery. We repeated the analysis after exclusion of two patients with intracranial surgery.

#### Results

After removal of the two patients with intracranial surgery, results remained unchanged for preoperative functional connectivity: $B_{VTA\text{-}PC4}$=-0.194 [-0.556; -0.123], $OR_{VTA\text{-}PC4}$=0.759 [0.573; 0.885], $p_{VTA\text{-}PC4}$=0.0057 and $B_{SNC\text{-}PC1}$=0.165 [0.028; 0.389], $OR_{SNC\text{-}PC1}$=1.180 [1.029; 1.476], $p_{SNC\text{-}PC1}$=0.0106, on 200/213 degrees of freedom.

#### Sex-specific analyses

##### Rationale and approach

Analyses of preoperative functional connectivity were repeated after stratification for sex. Model specifications were identical to those described in the main manuscript.

### Results

**Supplementary table S4:** Associations of preoperative VTA functional connectivity with POCD for female and male patients separately.

| | **Female patients** | | | **Male patients** | | |
|---|---|---|---|---|---|---|
| | **B (95% CI)** | **OR (95% CI)** | **p#** | **B (95% CI)** | **OR (95% CI)** | **p†** |
| **PC1** | -0.014 (-13.426; 11.197) | 0.986 (<0.001; >1000) | 0.9 | 0.007 (-0.754; 0.722) | 1.007 (0.470; 2.060) | 0.9 |
| | -0.014‡ (-12.551; 10.899) | 0.986‡ (<0.001; >1000) | 0.9‡ | 0.007‡ (-0.724; 0.700) | 1.007‡ (0.485; 2.014) | 0.9‡ |
| **PC2** | -0.196 (-3.171; 5.803) | 0.821 (<0.001; 331) | 0.19 | 0.156 (-0.141; 3.295) | 1.169 (0.868; 26.988) | 0.13 |
| | -0.196‡ (-28.709; 5.012) | 0.822‡ (<0.001; 150) | 0.19‡ | 0.156‡ (-0.142; 3.262) | 1.169‡ (0.868; 26.090) | 0.13‡ |
| **PC3** | -0.076 (-29.975; 8.097) | 0.927 (<0.001; >1000) | 0.5 | -0.048 (-0.966; 0.379) | 0.953 (0.381; 1.461) | 0.6 |
| | -0.076‡ (-28.406; 7.413) | 0.927‡ (<0.001; >1000) | 0.5‡ | -0.048‡ (-0.987; 0.370) | 0.953‡ (0.373; 1.448) | 0.6‡ |
| **PC4** | -0.378 (-70.715; 0.508) | 0.685 (<0.001; 1.662) | 0.055 | -0.257 (-7.397; 0.003) | 0.773 (0.001; 1.003) | 0.058 |
| | -0.378 ‡ (-68.569; 0.485) | 0.685‡ (<0.001; 1.624) | 0.056‡ | -0.257‡ (-8.177; 0.002) | 0.773‡ (<0.001; 1.002) | 0.058‡ |

### calculated on 73/85 degrees of freedom

† calculated on 115/127 degrees of freedom

‡ Results after exclusion of patients with intracranial surgery (one man, one woman), yielding 73/84 (women) and 115/126 (men) degrees of freedom

**Supplementary table S5:** Associations of preoperative SNc functional connectivity with POCD for female and male patients separately.

| | Female patients | | | Male patients | | |
|---|---|---|---|---|---|---|
| | **B (95% CI)** | **OR (95% CI)** | **p[#]** | **B (95% CI)** | **OR (95% CI)** | **p** |
| **PC1** | 0.337 (-0.106; 158.5) | 1.401 (0.899; >1000) | 0.017 | 0.128 (-0.137; 1.517) | 1.136 (0.872; 4.559) | 0.19 |
| | 0.337[‡] (-0.008; 155.9) | 1.401[‡] (0.920; >1000) | 0.017[‡] | 0.128[‡] (-0.134; 1.622) | 1.136[‡] (0.875; 5.064) | 0.19[‡] |
| **PC2** | 0.103 (-23.44; 48.86) | 1.109 (<0.001; >1000) | 0.45 | -0.004 (-0.428; 0.571) | 0.996 (0.651; 1.769) | >0.9 |
| | 0.103[‡] (-2.387; 46.43) | 1.109[‡] (<0.001; >1000) | 0.45[‡] | -0.004[‡] (-0.426; 0.574) | 0.996[‡] (0.653; 1.775) | >0.9[‡] |
| **PC3** | -0.043 (-72.680; 50.471) | 0.957 (<0.001; >1000) | 0.8 | -0.120 (-1.098; 0.113) | 0.887 (0.333; 1.119) | 0.24 |
| | -0.043[‡] (-0.717; 49.07) | 0.957[‡] (<0.001; >1000) | 0.8[‡] | -0.120[‡] (-1.127; 0.109) | 0.887[‡] (0.324; 1.115) | 0.24[‡] |
| **PC4** | -0.361 (-1.894; 2.343) | 0.697 (<0.001; 10.041) | 0.098 | 0.107 (-0.315; 1.147) | 1.113 (0.730; 3.148) | 0.42 |
| | -0.361[‡] (-0.019; 1.827) | 0.697[‡] (<0.001; 6.218) | 0.098[‡] | 0.107[‡] (-0.309; 1.132) | 1.113[‡] (0.735; 3.103) | 0.42[‡] |

[#] calculated on 73/85 degrees of freedom

[†] calculated on 115/127 degrees of freedom

[‡] Results after exclusion of patients with intracranial surgery (one man, one woman), yielding 73/84 (women) and 115/126 (men) degrees of freedom

79. Brooks JCW, Zambreanu L, Godinez A, Craig ADB, Tracey I. Somatotopic organisation of the human insula to painful heat studied with high resolution functional imaging. Neuroimage

2005;27(1):201–9.
